# Comparisons of Proprotein Convertase Subtilisin/Kexin Type 9 Inhibitors (PCSK9I) versus Ezetimibe on Major Adverse Cardiovascular Events Amongst Patients with Dyslipidaemia: A Population-Based Study

**DOI:** 10.1101/2023.09.23.23296003

**Authors:** Oscar Hou In Chou, Lifang Li, Cheuk To Skylar Chung, Lei Lu, Quinncy Lee, Hugo Hok Him Pui, Bosco Kwok Hei Leung, Carlin Chang, Tong Liu, Abraham Ka Chung Wai, Gregory Lip, Bernard Man Yung Cheung, Gary Tse, Jiandong Zhou

**Author notes:** Correspondence to: Gary Tse, MD DM PhD FRCP, Kent and Medway Medical School, Canterbury, United Kingdom, Tianjin Institute of Cardiology, The Second Hospital of Tianjin Medical University, Tianjin 300211, China, School of Nursing and Health Studies, Hong Kong Metropolitan University, Hong Kong, China, Jiandong Zhou, PhD, Division of Health Science, Warwick Medical School, University of Warwick, United Kingdom. Contributed equally (Oscar Hou In Chou, Lifang Li).

## Abstract

**Background:** Proprotein convertase subtilisin/kexin type 9 inhibitors (PCSK9I) have potential benefits against cardiovascular disease. The comparative risks of new-onset major adverse cardiovascular events (MACE) between PCSK9I and ezetimibe remain unknown.

**Objective:** This real-world study compared the risks of MACE upon exposure to PCSK9I and ezetimibe.

**Methods:** This was a retrospective population-based cohort study of patients with dyslipidaemia on either PCSK9I or ezetimibe between 1^st^ January 2015 and 30^th^ October 2022 using a territory-wide database from Hong Kong. The primary outcome was new-onset MACE. The secondary outcomes were myocardial infarction, heart failure, stroke/transient ischaemic attack, and all-cause mortality. Propensity score matching (1:3 ratio) using the nearest neighbour search was performed. Multivariable Cox regression was used to identify significant associations.

**Results:** This cohort included 42450 dyslipidaemia patients (median age: 65.0 years old [SD: 11.1]; 64.54 % males). The PCSK9I and ezetimibe groups consisted of 1477 and 40973 patients, respectively. After matching, 67 and 235 patients suffered from MACE in the PCSK9I and ezetimibe groups, respectively, over a total of 14514.5 person-years. PCSK9I was associated with lower risks of MACE (Hazard ratio [HR]: 0.59; 95% Confidence Interval [CI]: 0.37-0.92) compared to ezetimibe use after adjusting for demographics, past comorbidities, other medications, and time-weighted means of lipid and glucose tests. Besides, while both alirocumab and evolocumab were associated with lower risks of MACE, evolocumab was associated with significantly lower risks of myocardial infarction, heart failure, and stroke/transient ischaemic attack. The results remained consistent in the competing risk and sensitivity analyses.

**Conclusions:** PCSK9I use amongst dyslipidaemia patients was associated with lower risks of new-onset MACE and outcomes compared to ezetimibe after adjustments. Evolocumab might perform better than Alirocumab in reducing the risks of cardiovascular diseases.

**Illustrated Abstract:** 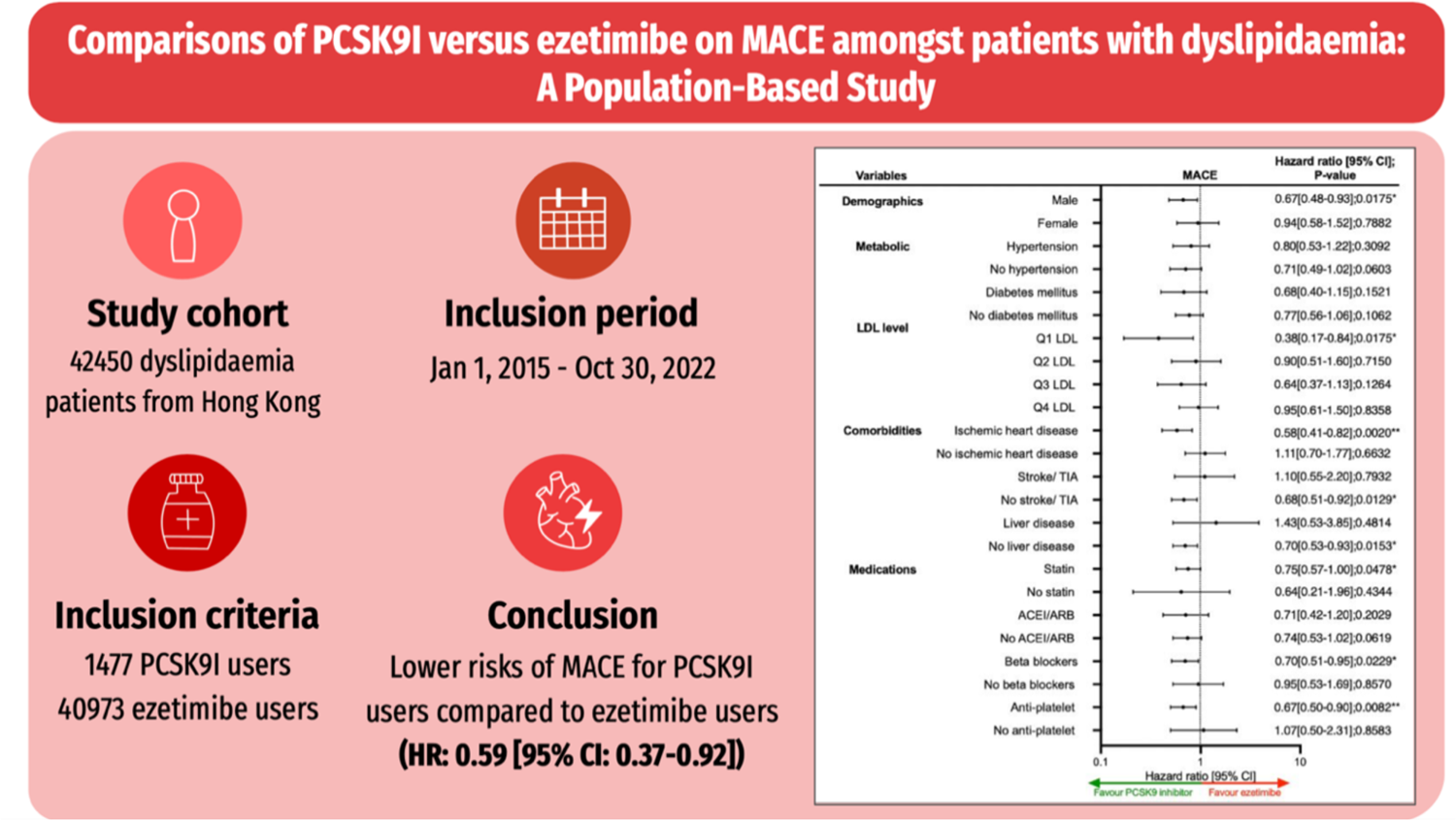

## Introduction

The growing prevalence of dyslipidaemia continues to impose a burden on healthcare systems worldwide ^1^, by increasing the risks of atherosclerosis ^2^ and adverse cardiovascular outcomes ^3, 4^. Statins are prescribed as a first-line therapy to improve cholesterol management and cardiovascular outcomes amongst patients with dyslipidaemia.^5, 6^ However, this preventive effect is only applicable for low-risk patients.^7^ To optimise treatment for cardiovascular disease prevention amongst patients insufficient controlled with statin, this garnered interests in second-line lipid-lowering agents such as proprotein convertase subtilisin/kexin type 9 inhibitors (PCSK9I) and ezetimibe on major adverse cardiovascular events (MACE) outcomes.^8^ The association between these lipid-lowering drugs and atherosclerotic cardiovascular diseases have been explored.^9, 10^ Recent study highlight that PCSK9I reduce the occurrence of MACE more effectively than ezetimibe when supplemented into background therapy for high-risk patients.^11^ For Instance, the FOURIER (Further Cardiovascular Outcomes Research with PCSK9 Inhibition subjects with Elevated Risk) study showed evidence that evolocumab usage reduced cardiovascular outcomes and mortality compared to placebo.^12^

While PCSK9I had demonstrated a significant reduction in low-density lipoprotein (LDL) levels and cardiovascular outcomes, the cost of PCSK9 inhibitors raised concern about the efficacy of the drug relative to its cost.^13^ It is imperative to compare whether these second-line lipid-lowering agents reduce the risk of MACE. A simulation model predicts that combination therapy of statin with each respective drug can successfully achieve a relative reduction of 5-6% in the next five years, with the highest predicted absolute aversion of MACE occurring in China.^14^ Previously, there was sparse information in guidelines that assess the effects of PCSK9I and ezetimibe on the primary and secondary prevention of cardiovascular disease across all risk groups.^7, 11^ To corroborate, there is a need of more clinical data on the head-to-head comparison between PCSK9I or ezetimibe and MACE. An observational study suggests that PCSK9I users had improved arterial elasticity compared to ezetimibe.^15^ Thus, this study aimed to explore the role of PSCK9I and ezetimibe with new-onset MACE and other cardiovascular outcomes in a territory-wide cohort study.

## Methods

This study was approved by the Institutional Review Board of the University of Hong Kong/Hospital Authority Hong Kong West Cluster Institutional Review Board (HKU/HA HKWC IRB) (UW-20-250 and UW 23-339) and complied with the Declaration of Helsinki.

### Study population

This was a retrospective population-based study of prospectively collected electronic health records using the Clinical Data Analysis and Reporting System (CDARS) by the Hospital Authority (HA) of Hong Kong. These records include information from various healthcare facilities in Hong Kong, such as hospitals, clinics, and day-care centres. This system has been used extensively by our teams and other research teams in Hong Kong ^16, 17^. The study focused on patients with dyslipidaemia who were given PCSK9I or ezetimibe treatment in HA centres from January 2015 to October 2022.

### Predictors and variables

Patients’ demographics include gender and age of initial drug use (baseline), and clinical and biochemical data were extracted for the present study. Prior comorbidities were extracted by the International Classification of Diseases Ninth Edition (ICD-9) codes (**Supplementary Table 1**). The level of the LDL was calculated using Friedewald formula from total cholesterol and high-density lipoprotein (HDL). Dyslipidaemia duration was calculated by examining the earliest date amongst the first date of (1) diagnosis using ICD-9; (2) Total cholesterol >5.2 mmol/l or LDL cholesterol >3.4 mmol/l or HDL cholesterol <1 mmol/l in men and <1.3 mmol/L in women; (3) using anti-lipid medications. The patients on financial aid were defined as patients on the Comprehensive Social Security Assistance (CSSA) scheme, higher disability allowance, normal disability allowance, waiver, and other financial aid in Hong Kong. The number of hospitalisations in the year prior to the index days was extracted. The Charlson standard comorbidity index was calculated.^18^ The duration and frequency of PCSK9I and ezetimibe usage was calculated. The baseline laboratory examinations, including the glucose profiles and renal function tests, were extracted. The estimated glomerular filtration rate (eGFR) was calculated using the abbreviated modification of diet in renal disease (MDRD) formula.^19^ The time-weighted lipid and glucose profiles were also calculated by the products of the sums of two consecutive measurements and the time interval, then divided by the total time interval, as suggested previously.^20^

### Study outcomes

The primary outcome of this study was new-onset MACE defined by a composite of cardiovascular mortality, myocardial infarction, heart failure, and stroke using International Classification of Diseases Ninth Edition (ICD-9) codes (**Supplementary Table 1)** according to the verified published literature.^21^ The secondary outcomes were myocardial infarction, heart failure, stroke/transient ischaemic attack, and all-cause mortality. Mortality data were obtained from the Hong Kong Death Registry, a population-based official government registry with the registered death records of all Hong Kong citizens linked to CDARS. Mortality was recorded using the *International Classification of Diseases Tenth Edition* (ICD-10) coding. The as-treat approach was adopted which patients were censored at treatment discontinuation or switching of the comparison medications. The endpoint date of interest for eligible patients was the event presentation date. The endpoint for those without primary outcome was the mortality date or the end of the study period (30^th^ April 2023).

### Statistical analysis

Descriptive statistics were used to summarize baseline clinical and biochemical characteristics of patients with PCSK9I and ezetimibe use. For baseline clinical characteristics, continuous variables were presented as mean (95% confidence interval/standard deviation), and the categorical variables were presented as total numbers (percentage). Propensity score matching generated by logistic regression with a 1:3 ratio for PCSK9I use versus ezetimibe use based on demographics, past comorbidities, non-PCSK9I /ezetimibe medications, duration from hyperlipidaemia to index date, MDRD for eGFR, number of hospitalisation, average episode stay length, number of anti-hypertensive drugs, number of anti-diabetic drug classes, and time-weighted lipid and glucose tests were performed using the nearest neighbour search strategy with a calliper of 0.1. Propensity score matching was performed using Stata software (Version 16.0). Baseline characteristics between patients with PCSK9I and ezetimibe use before and after matching were compared using the absolute standardized mean difference (SMD), with SMD<0.20 regarded as well-balanced between the two groups.

The cumulative incidence curves for the primary outcomes and secondary outcomes were constructed. Cox regression was used to identify significant risk predictors of adverse study outcomes in the matched cohort, with adjustments for significant demographics, past comorbidities, duration from hyperlipidaemia, and number of prior hospitalizations, non-PCSK9i or Ezetimibe medications, abbreviated MDRD, and time-weighted means of lipid and glucose tests. The log-log plot was used to verify the proportionality assumption for the proportional Cox regression models. Subgroup analyses were conducted to confirm the association amongst patients with different clinically significant predictors accounting for dyslipidaemia, as well as the comorbidities and medications associated with MACE.

Cause-specific and sub-distribution hazard models were conducted to consider possible competing risks. Multiple approaches using the propensity score were used, including propensity score stratification,^22^ propensity score with inverse probability of treatment weighting (IPTW) ^23^ and propensity score with stable inverse probability weighting.^24^ Sensitivity analyses result with consideration of patients with or without prior MACE, patients with less than 3-month follow-up duration or drug exposure, patients who died within 30 days after drug exposure, patients with liver dysfunction, and patients at the top or bottom 5% of propensity score matching. We used the hip fracture as the negative control in the falsification analysis **(Supplementary Table 1)**, such that the observed significant association in the falsification analysis should be attributed to bias. The hazard ratio (HR), 95% CI, and P-value were reported. Statistical significance was defined as P-value <0.05. All statistical analyses were performed with RStudio (Version: 1.1.456) and Python (Version: 3.6).

## Results

In this territory-wide cohort study of 42450 patients with dyslipidaemia treated with PCSK9I or ezetimibe between 1^st^ January 2015 and 30^th^ October 2022 in Hong Kong, patients were followed up until 30^th^ April 2023 or until their deaths (**Figure 1**). This study included a total of 42450 patients with dyslipidaemia (mean age: 65.0 years old [SD: 11.1]; 64.54% males), of whom 1477 patients used PCSK9I, and 40973 patients used ezetimibe (**Table 1**). Before matching, the PCSK9I users were younger, with fewer comorbidities, with more cardiovascular diseases (ischaemic heart disease), using more anti-lipid drugs, more anti-platelet drugs, higher low-density lipoprotein, and higher total cholesterol.

**Figure 1.**
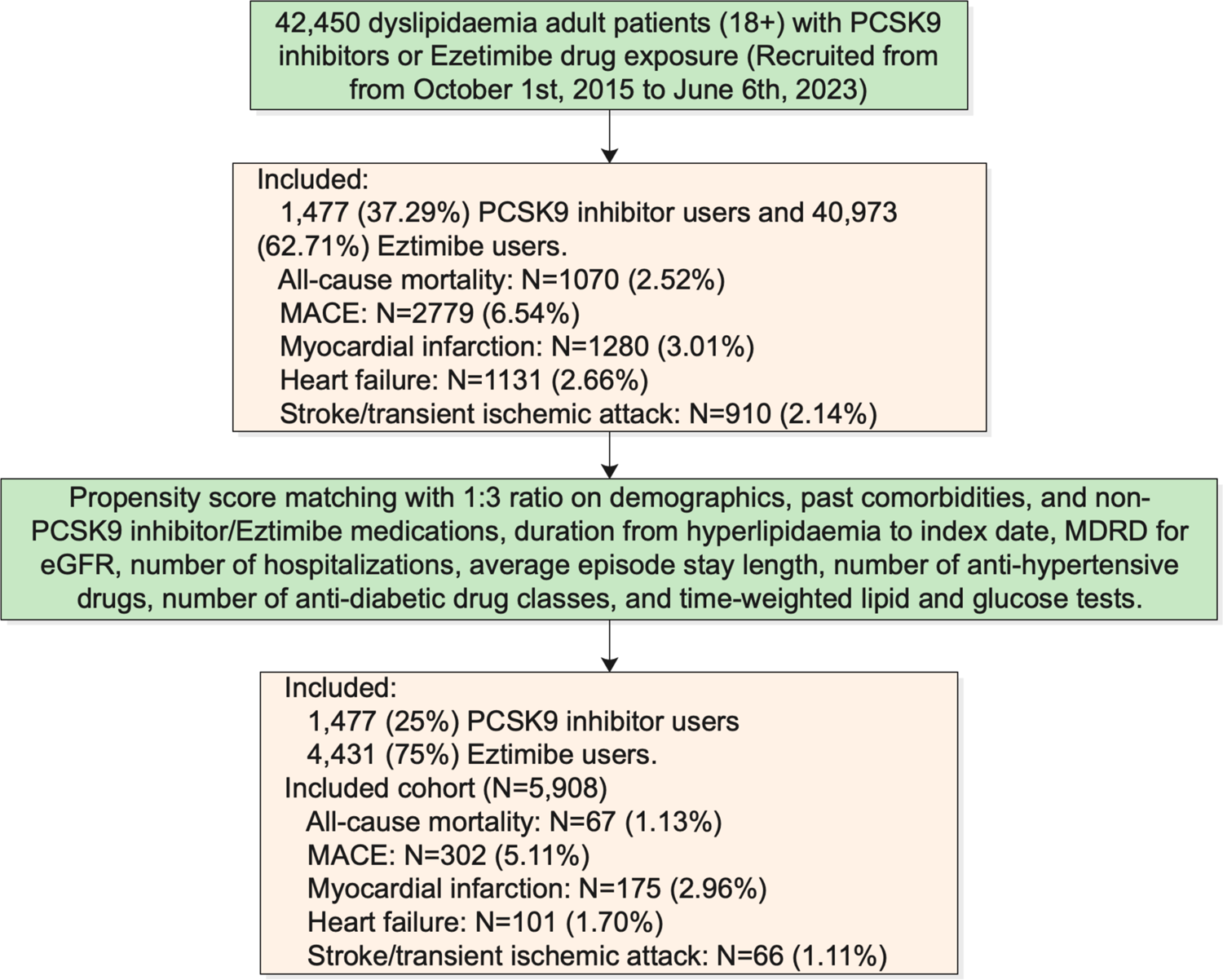
Procedures of data processing for the study cohort PCSK9: Proprotein convertase subtilisin kexin 9; MDRD: modification of diet in renal disease

**Table 1.**
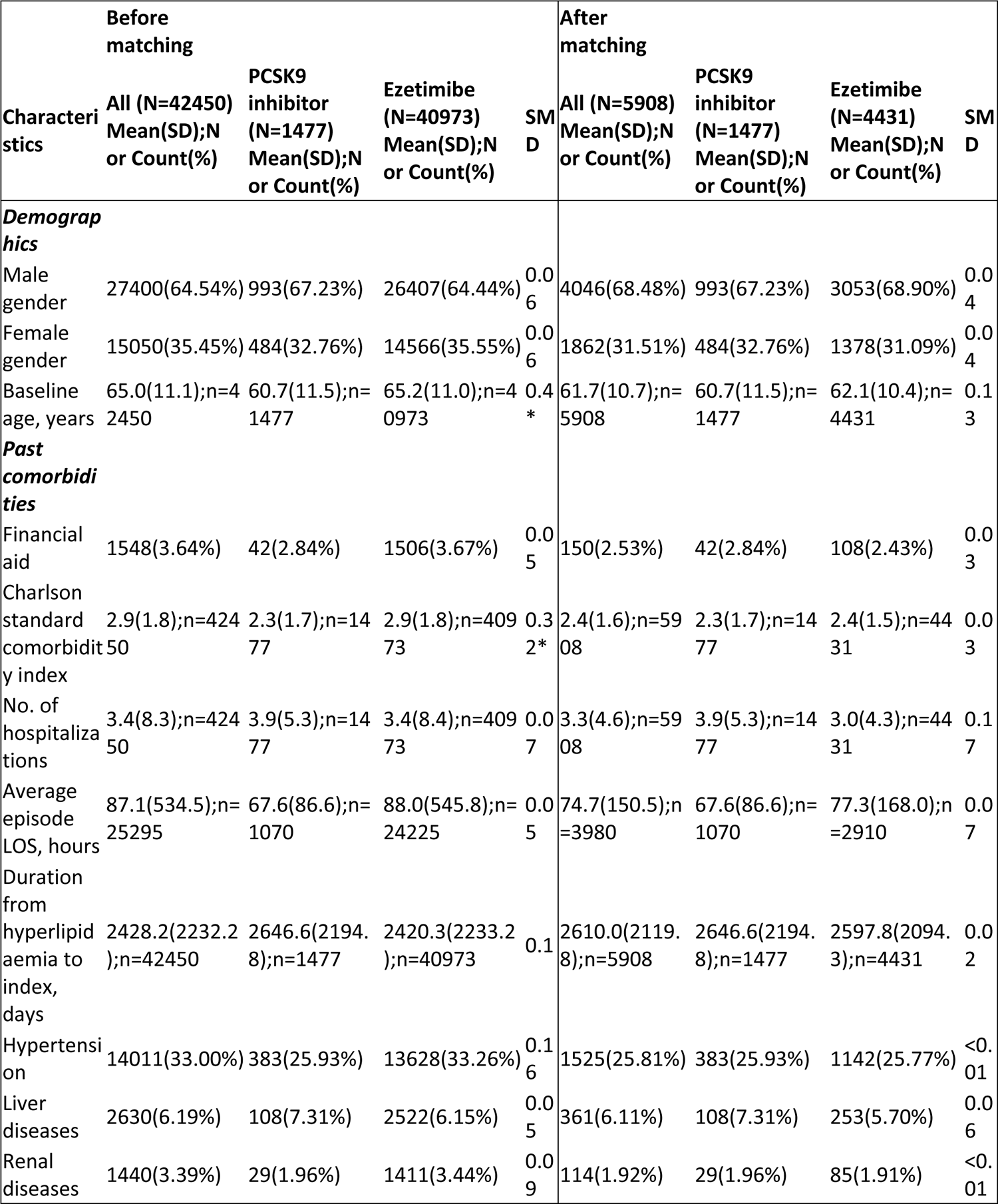
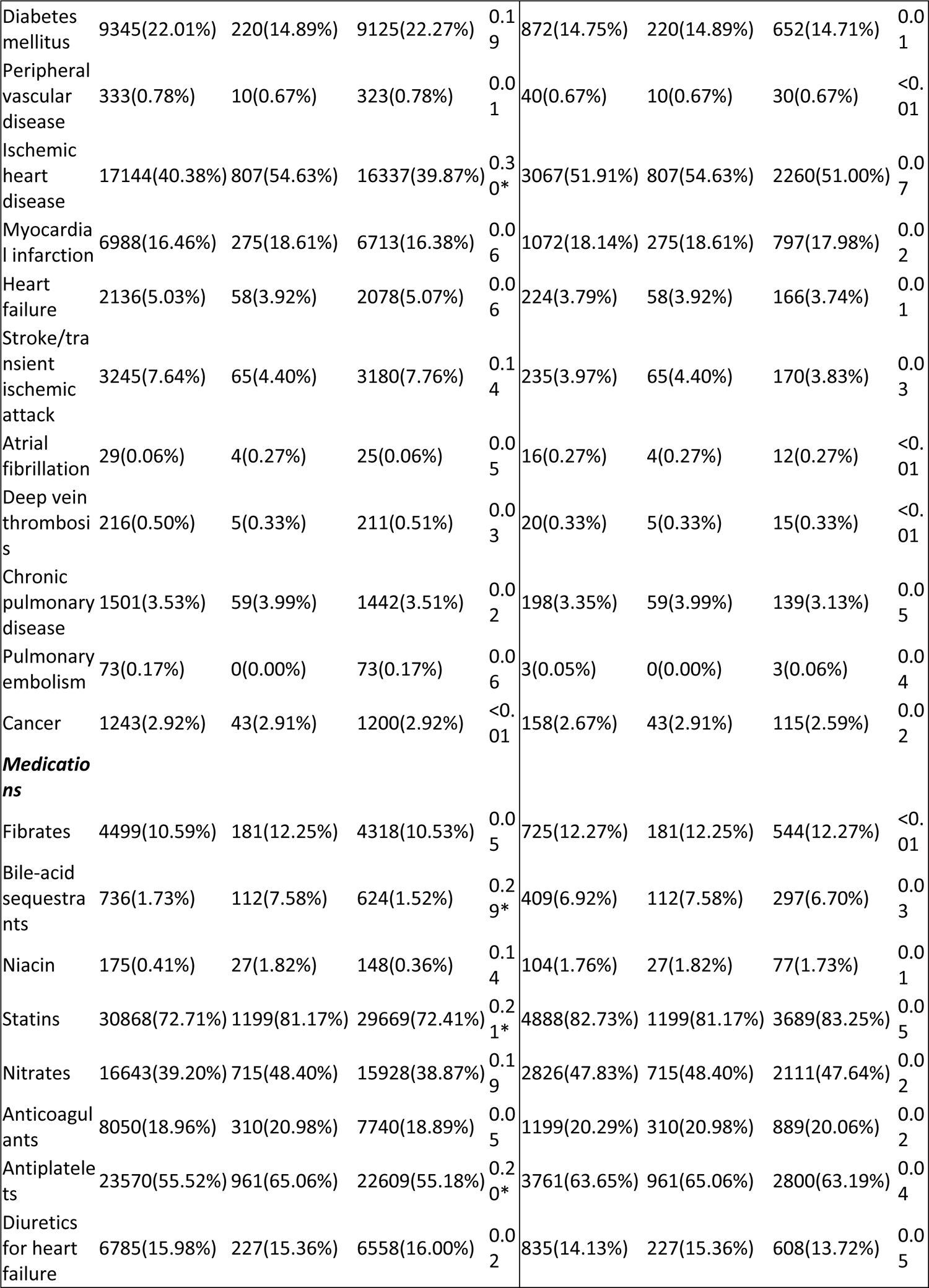
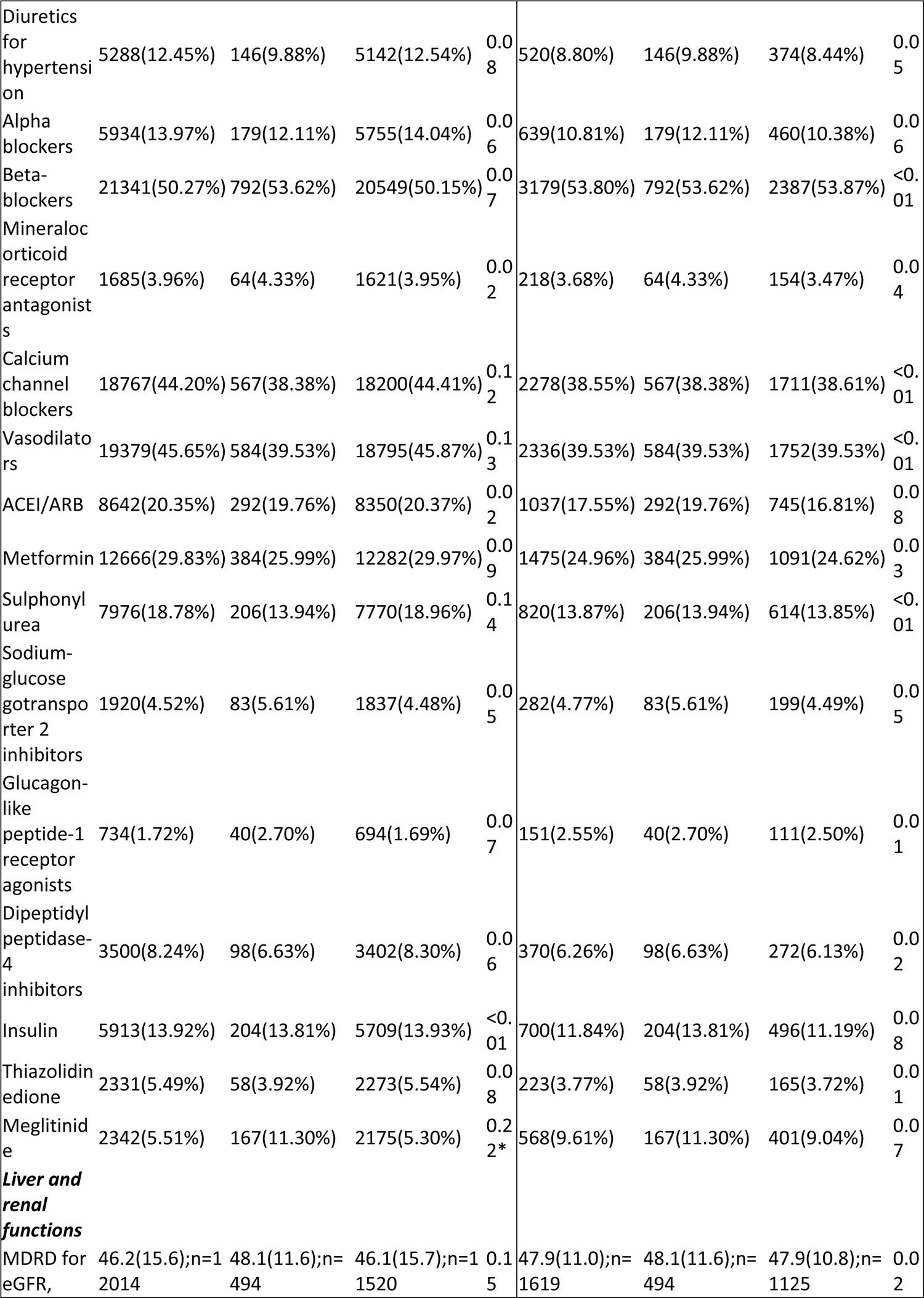
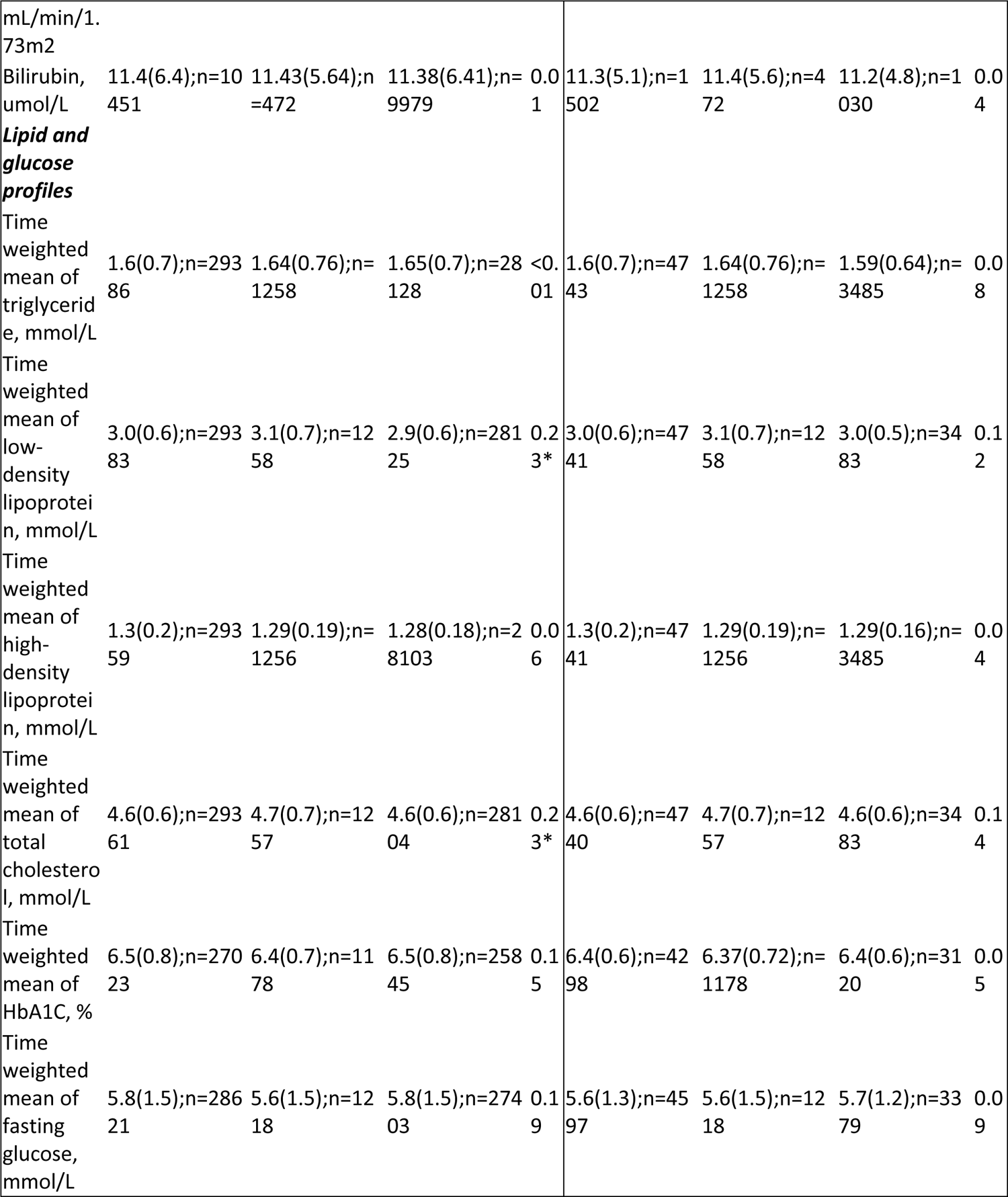
Baseline and clinical characteristics of patients with PCSK9 inhibitor v.s. Ezetimibe use before and after propensity score matching (1:3). * for SMD≥0.2; PCSK9: Proprotein convertase subtilisin kexin 9; SD: standard deviation; CV: Coefficient of variation; MDRD: modification of diet in renal disease; PCSK9: Proprotein convertase subtilisin kexin 9; ACEI: angiotensin-converting enzyme inhibitors; ARB: angiotensin II receptor blockers.

After the propensity score matching, baseline characteristics and the time-weighted lipid and glucose profiles of the two groups were well-balanced, apart from the SD of total cholesterol (**Table 1**). The PCSK9I and ezetimibe cohorts were comparable after matching with the nearest neighbour search strategy with a calliper of 0.1, and the proportional hazard assumption was confirmed (**Supplementary Figure 1**). In the matched cohort, 302 patients developed MACE; 67 patients passed away during the study period (**Figure 1**). The characteristics of patients are shown in **Table 1**.

### Association between PCSK9I and ezetimibe and MACE

In the matched cohort, 67 PCSK9I users and 216 ezetimibe users developed MACE. After a mean follow-up of 14514.5 person-year, the incidence of MACE was lower amongst PCSK9I users (Incidence rate [IR] per 1000 person-year: 14.96; 95% CI: 11.60-19.01) compared to ezetimibe users (IR per 1000 person-year: 21.52; 95% CI: 18.74-24.59) (**Table 2**). PCSK9I users had a 41% lower risk of MACE after adjustment (HR: 0.59; 95% CI: 00.37-0.92, p=0.0203) compared to users of ezetimibe users regardless of the demographics, comorbidities, medication profile, renal function, glycaemic tests, number of hospitalisations, and the duration of dyslipidaemia (**Table 2; Supplementary Table 2)**. This was substantiated by the cumulative incidence curves stratified by PCSK9 versus ezetimibe (**Figure 2**).

**Figure 2.**
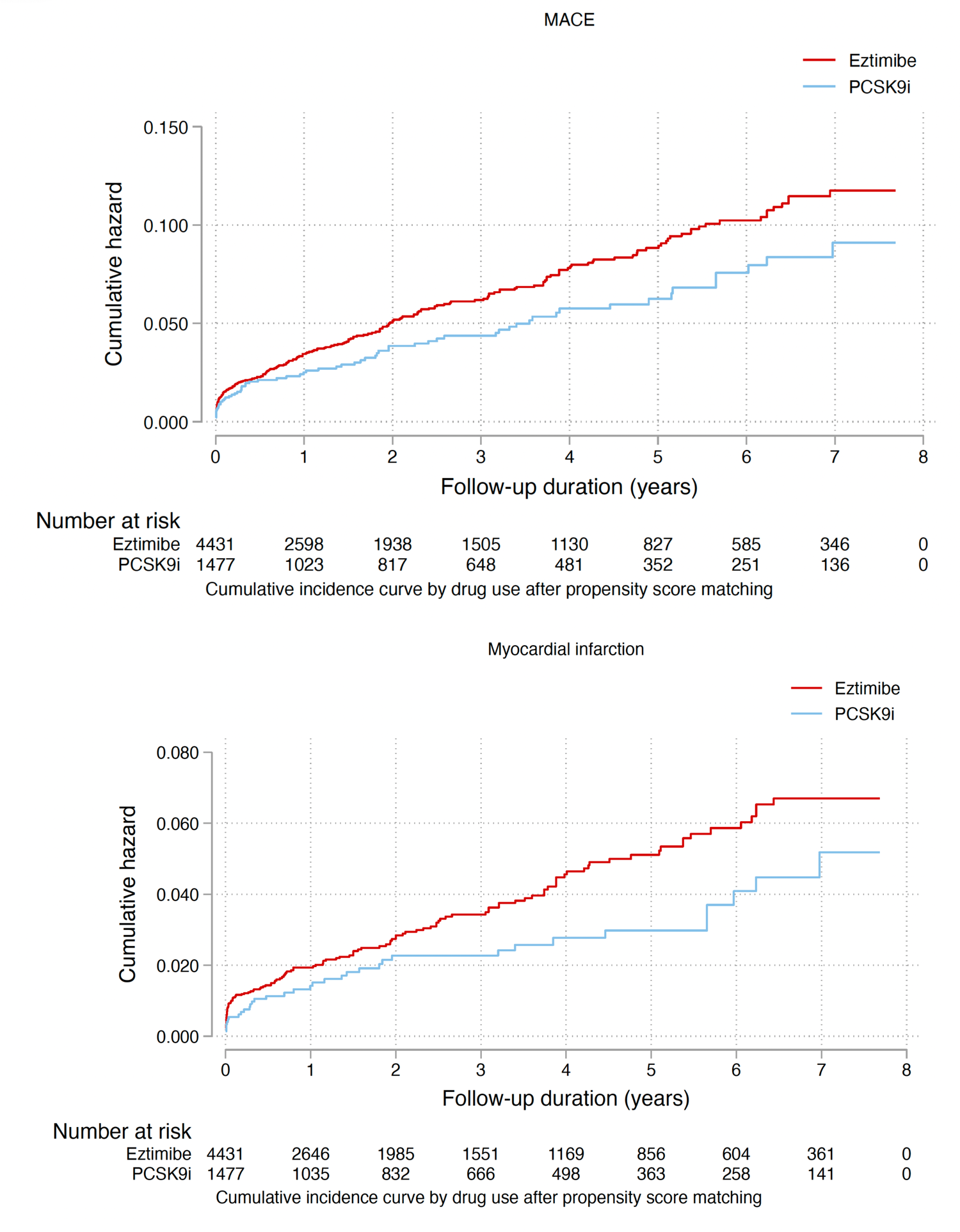
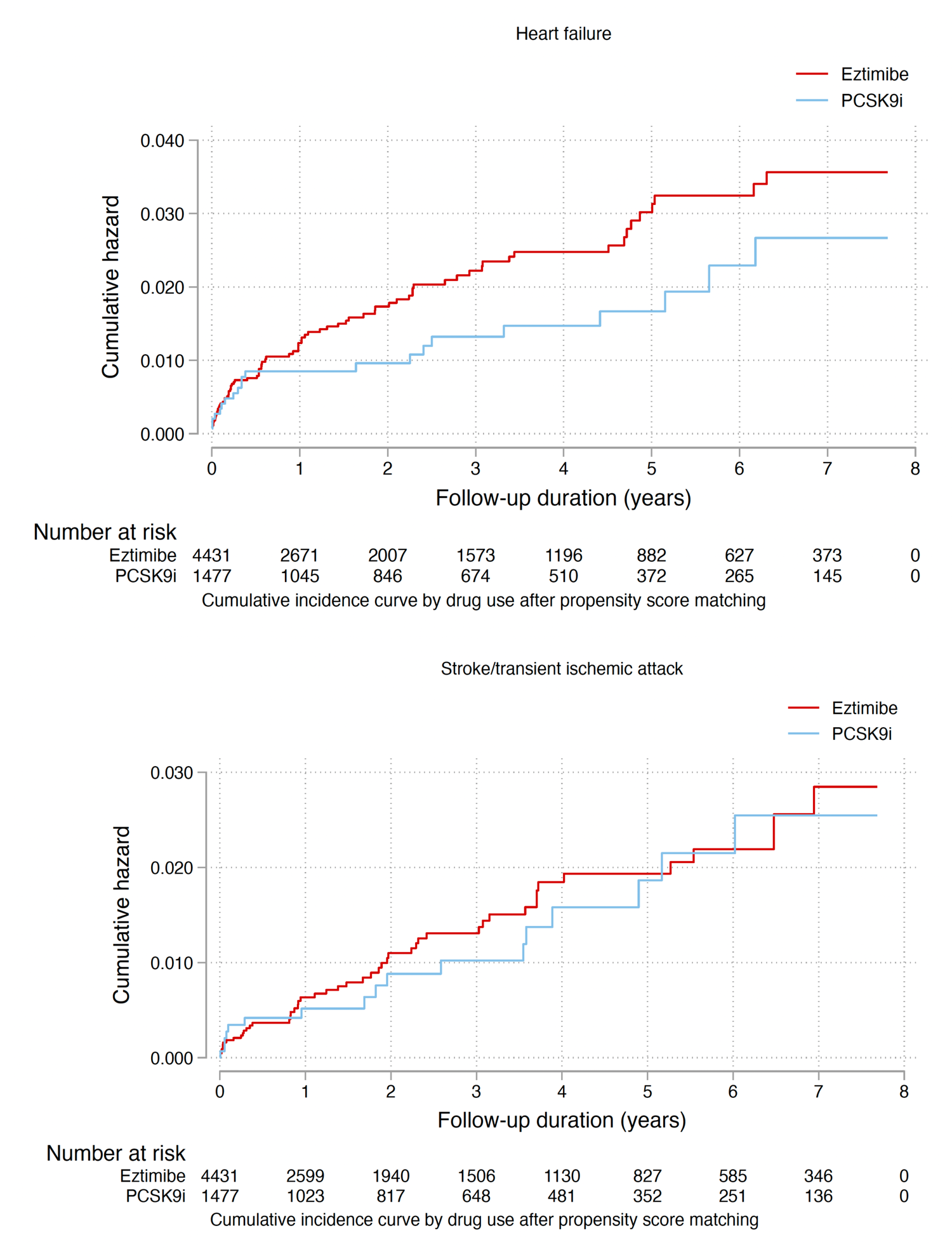
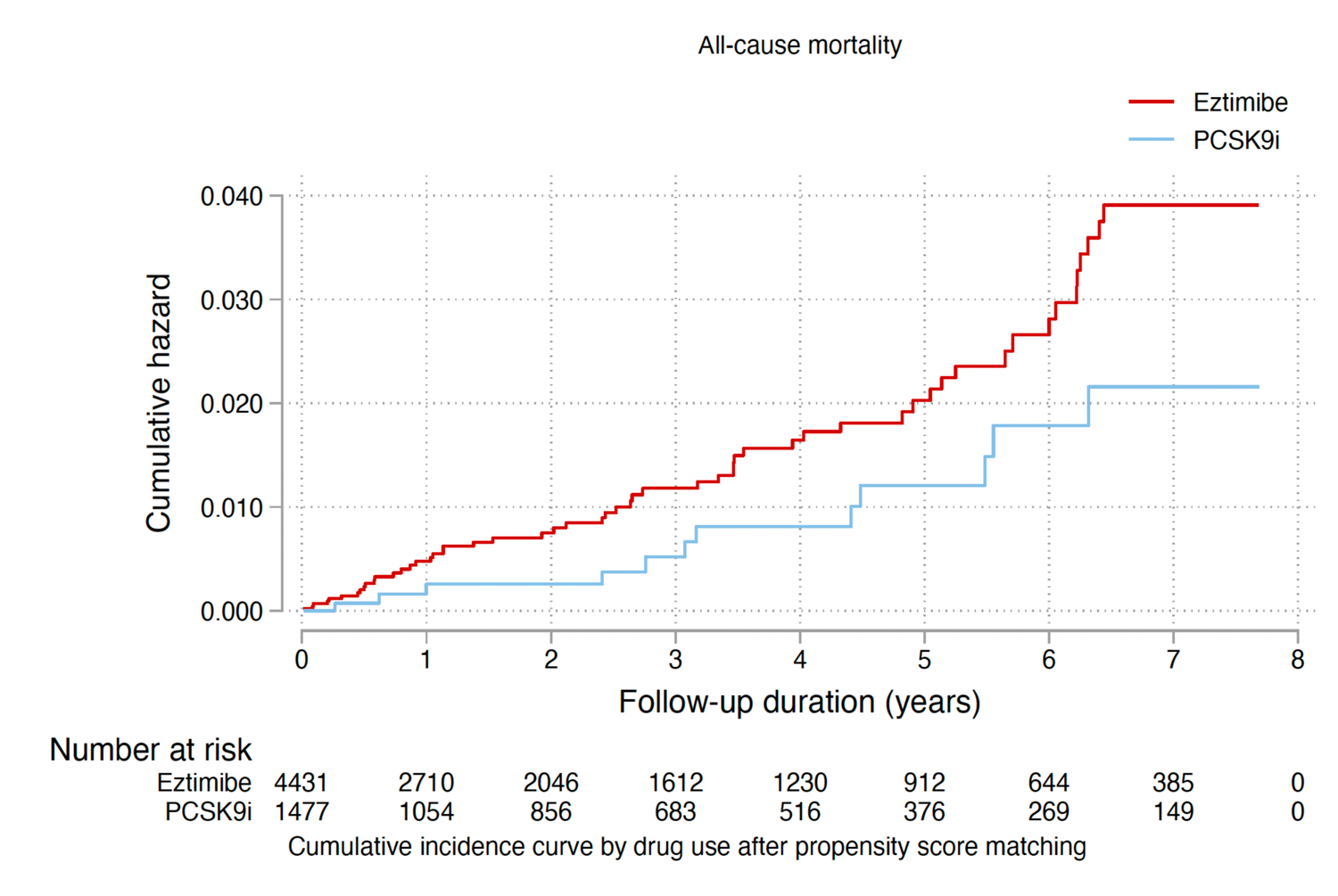
Cumulative incidence curves for new onset MACE, myocardial infarction, heart failure, stroke/transient ischemic attack, and all-cause mortality stratified by drug exposure effects of PCSK9 inhibitor and ezetimibe after propensity score matching (1:3). PCSK9: Proprotein convertase subtilisin kexin 9;

**Table 2.** Incidence rate (IR) per 1000 person-year and multivariate Cox regression models of new onset MACE and all-cause mortality in the cohort after 1:3 propensity score matching. * for p≤ 0.05, ** for p ≤ 0.01, *** for p ≤ 0.001; CI: confidence interval; PCSK9: Proprotein convertase subtilisin kexin 9. Adjusted for significant demographics, past comorbidities, duration from hyperlipidaemia, and number of prior hospitalizations, number of anti-diabetic drugs, number of anti-hypertensive drugs, non PCSK9i or Ezetimibe medications, abbreviated MDRD, and time-weighted means of lipid and glucose tests.

| **MACE** | **Number of patients** | **Number of events** | **Total person-year** | **Incidence [95% CI] per 1000 person-year** | **Adjusted hazard ratio [95% CI]** | **P-value** |
| --- | --- | --- | --- | --- | --- | --- |
| **Ezetimibe** | 4431 | 235 | 10038.5 | 23.41[20.51-26.60] | 1 [Reference] | NA |
| **PCSK9 inhibitor** | 1477 | 67 | 4476 | 14.97[11.60-19.01] | 0.59[0.37-0.92] | 0.0203* |
| **Myocardial infarction** | **Number of patients** | **Number of events** | **Total person-year** | **Incidence [95% CI] per 1000 person-year** | **Adjusted hazard ratio [95% CI]** | **P value** |
| **Ezetimibe** | 4431 | 139 | 10267.6 | 13.54[11.38-15.99] | 1 [Reference] | NA |
| **PCSK9 inhibitor** | 1477 | 36 | 4567.3 | 7.88[5.52-10.91] | 0.55[0.28-1.08] | 0.0828 |
| **Heart failure** | **Number of patients** | **Number of events** | **Total person-year** | **Incidence [95% CI] per 1000 person-year** | **Adjusted hazard ratio [95% CI]** | **P value** |
| **Ezetimibe** | 4431 | 80 | 10628.7 | 7.52[5.97-9.37] | 1 [Reference] | NA |
| **PCSK9 inhibitor** | 1477 | 21 | 4682.3 | 4.48-2.78-6.86] | 0.89[0.46-1.76] | 0.7459 |
| **Stroke/transient ischemic attack** | **Number of patients** | **Number of events** | **Total person-year** | **Incidence [95% CI] per 1000 person-year** | **Adjusted hazard ratio [95% CI]** | **P value** |
| **Ezetimibe** | 4431 | 49 | 8268.4 | 5.93[4.38-7.83] | 1 [Reference] | NA |
| **PCSK9 inhibitor** | 1477 | 17 | 4476 | 3.80[2.21-6.08] | 0.50[0.18-1.41] | 0.1917 |
| **All-cause mortality** | **Number of patients** | **Number of events** | **Total person-year** | **Incidence [95% CI] per 1000 person-year** | **Adjusted hazard ratio [95% CI]** | **P value** |
| **Ezetimibe** | 1477 | 55 | 10249.2 | 5.36[4.04-6.98] | 1 [Reference] | NA |
| **PCSK9 inhibitor** | 4431 | 12 | 4630.8 | 0.26[1.34-4.53] | 0.45[0.17-1.20] | 0.1101 |

### Association between PCSK9I and ezetimibe and the secondary outcomes

After 1:3 propensity score matching, 36 PCSK9I users and 139 ezetimibe users developed myocardial infarction. PCSK9I use was not significantly associated with lower risk of myocardial infarction after adjustment (HR: 0.55; 95% CI: 0.28-1.08, p=0.0828) compared to users of ezetimibe (**Table 2**). Meanwhile, 21 PCSK9I users and 80 ezetimibe users developed heart failure. PCSK9I was not associated with lower risk of heart failure after adjustment (HR: 0.89; 95% CI: 0.46-1.76, p=0.7459) compared to users of ezetimibe (**Table 2; Supplementary Table 2)**. Furthermore, 17 PCSK9I users and 49 ezetimibe users developed stroke/transient ischaemic attack. PCSK9I was not associated with lower risk of stroke/transient ischaemic attack after adjustment (HR: 0.50; 95% CI: 0.18-1.41, p=0.1917) compared to users of ezetimibe (**Table 2; Supplementary Table 2)**. Overall, 12 PCSK9I users and 55 ezetimibe users passed away. PCSK9I was not associated with lower risk of all-cause mortality after adjustment (HR: 0.45; 95% CI: 0.17-1.20, p=0.1101) compared to users of ezetimibe (**Table 2; Supplementary Table 2)**.

### Relationship between alirocumab or evolocumab on the adverse outcomes

Amongst the users who only used alirocumab, alirocumab was associated with lower risks of MACE compared to ezetimibe (HR: 0.60; 95% CI: 0.38-0.98, p=0.0403). However, alirocumab was not associated with lower risks of myocardial infarction, heart failure, stroke/transient ischaemic attack and all-cause mortality compared to ezetimibe (all p>0.05). Meanwhile, amongst the users only used evolocumab, evolocumab not only was associated with lower risks of MACE compared to ezetimibe (HR: 0.15; 95% CI: 0.06-0.33, p<0.0001); evolocumab was also associated with lower risks of myocardial infarction, heart failure, stroke/transient ischaemic attack and all-cause mortality compared to ezetimibe (all p<0.05).

However, due to the limited power, evolocumab was not associated with a lower risk of MACE compared to alirocumab (HR: 0.49; 95% CI: 0.20-1.24, p=0.1323) (**Table 3**).

**Table 3.**
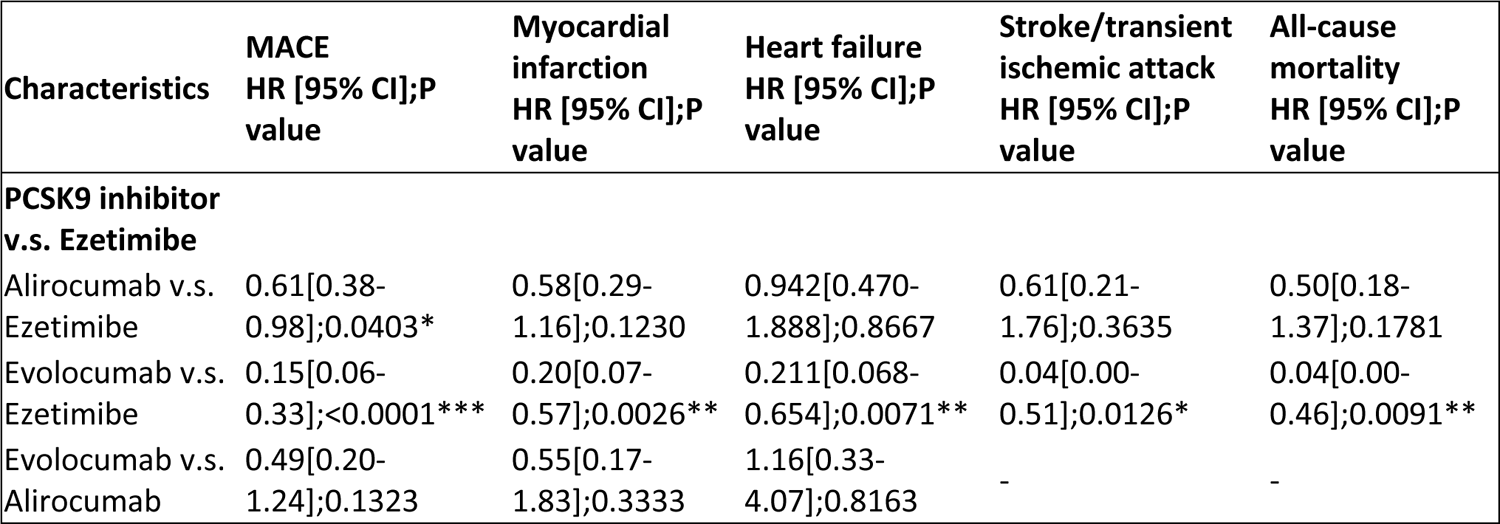
Multivariate Cox regression models of new onset MACE and all-cause mortality amongst Alirocumab or Evolocumab only users compared to Ezetimibe users in the cohort after 1:3 propensity score matching. * for p≤ 0.05, ** for p ≤ 0.01, *** for p ≤ 0.001; CI: confidence interval; PCSK9: Proprotein convertase subtilisin kexin 9. Adjusted for significant demographics, past comorbidities, duration from hyperlipidaemia, and number of prior hospitalizations, number of anti-diabetic drugs, number of anti-hypertensive drugs, non PCSK9i or Ezetimibe medications, abbreviated MDRD, and time-weighted means of lipid and glucose tests.

### Subgroup analysis

The results of the subgroup analysis for effects of PCSK9I and ezetimibe on the MACE are shown in **Figure 3**. The result demonstrated that PCSK9I was associated with lower risks of MACE than ezetimibe amongst male patients with Q1 LDL level, with prior history of ischaemic heart disease, without prior stroke/transient ischaemic attack, without prior liver disease, on beta-blocker and anti-platelets. Amongst patients who used only alirocumab, alirocumab was associated with lower risks of MACE than ezetimibe amongst patients with Q1 LDL level. Meanwhile, amongst patients who used only evolocumab, the results remained significant regardless of sex and hypertension. Furthermore, evolocumab was associated with lower risks of MACE than ezetimibe amongst patients with Q3 or Q4 LDL levels.

**Figure 3.**
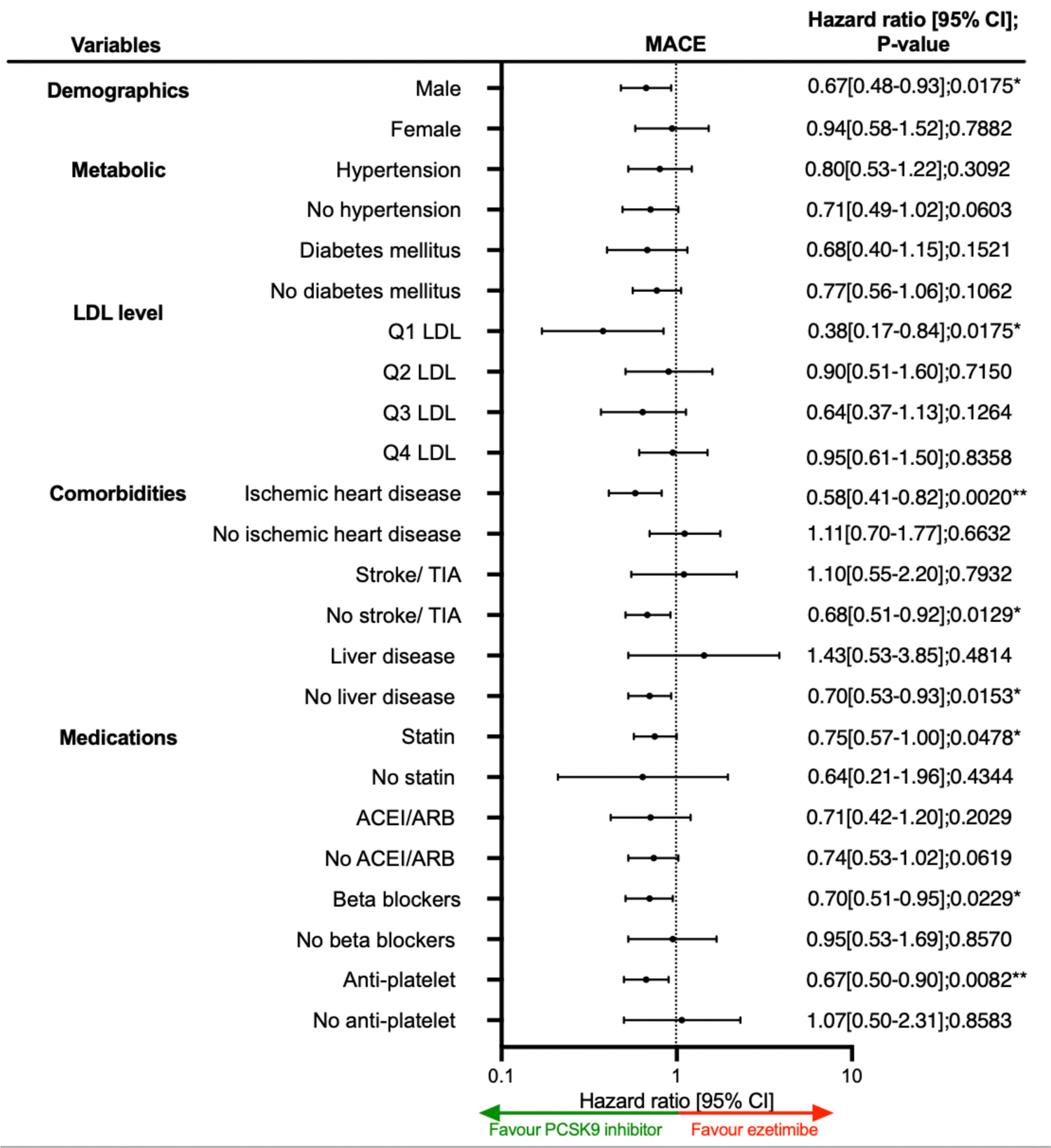
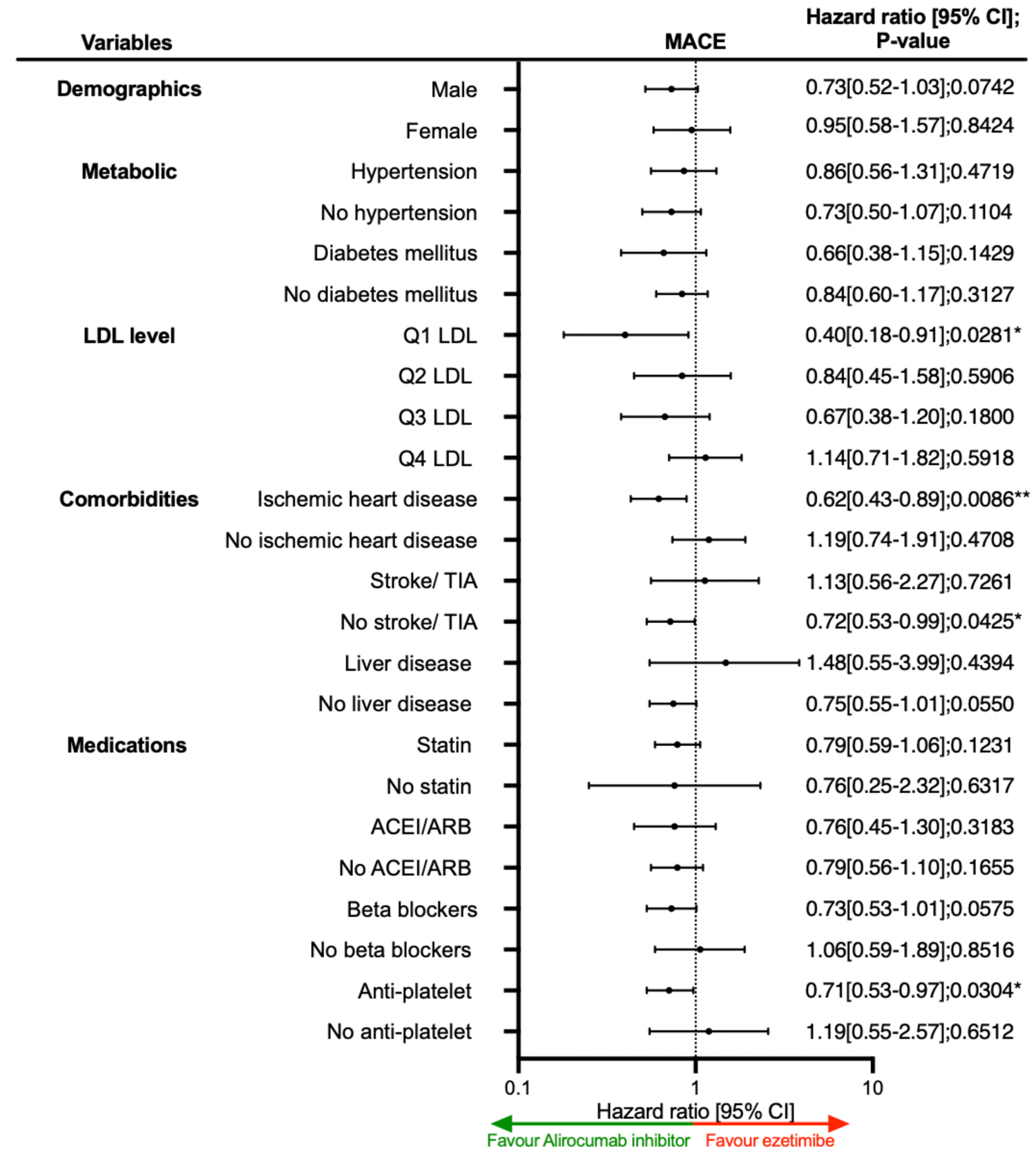
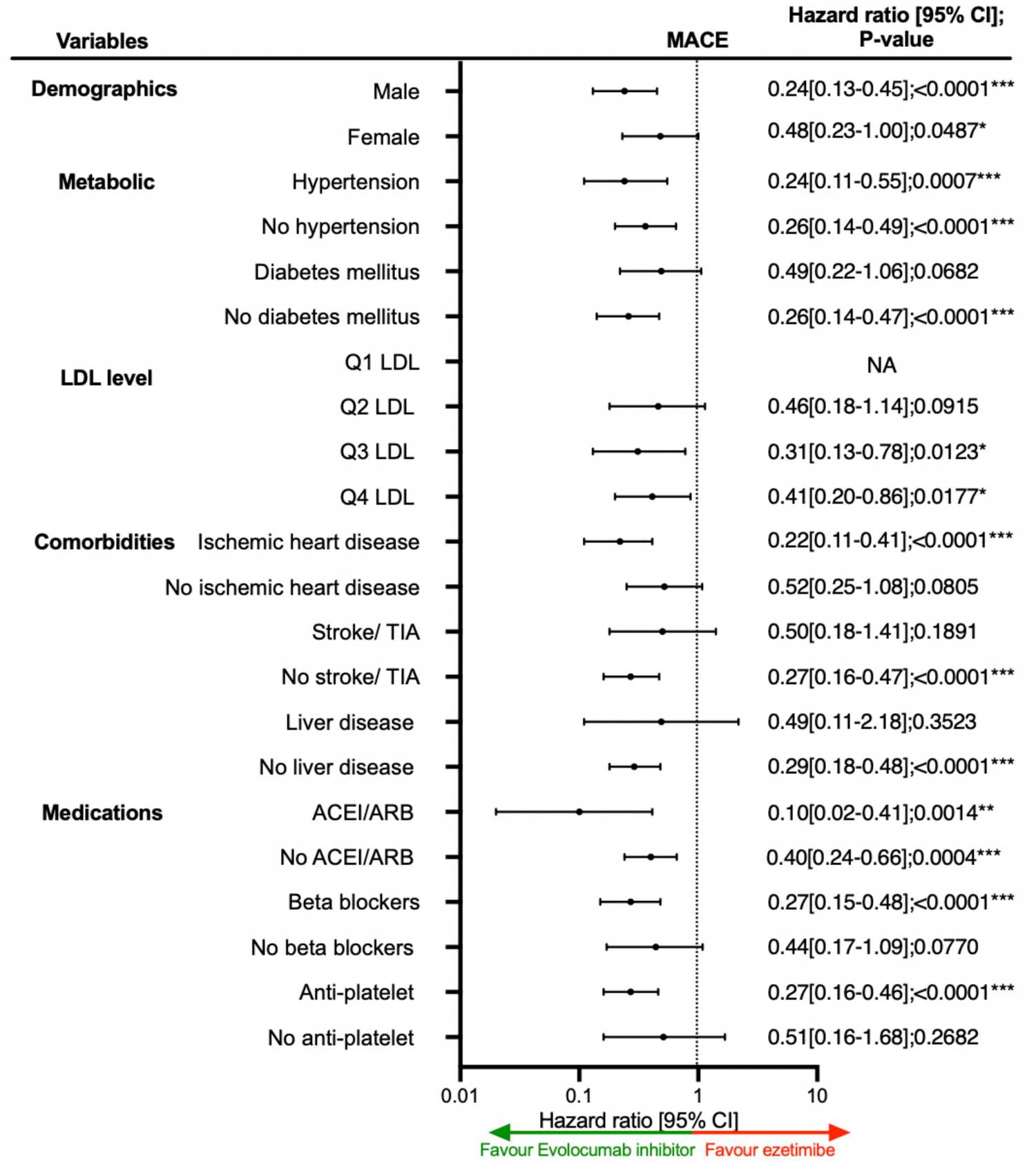
Forests plot of hazard ratios with 95% CI for PCSK9 inhibitor and its subtypes v.s. ezetimibe on new onset MACE in the matched cohort. (a) Between PCSK9 inhibitor and ezetimibe (b) Between alirocumab and ezetimibe (c) Between evolocumab and ezetimibe.

The marginal effects analysis demonstrated that PCSK9I was associated with lower risks of MACE amongst patients with lower levels of LDL; the differences between PCSK9I and ezetimibe narrowed as the level of LDL increased. The same trend was observed as well in myocardial infarction and heart failure. Meanwhile, for patients with LDL level <2 mmol/L, ezetimibe was associated with lower risks of stroke/transient ischaemic attack, and vice versa occurred as LDL level >2 mmol/L. Besides, PCSK9I was associated with lower risks of MACE and other cardiovascular outcomes as the duration of hyperlipidaemia and the number of lipid-lowering drugs increased (**Supplementary** Figure 3).

### Sensitivity analysis

Sensitivity analyses were performed to confirm the predictability of the models. The results of the cause-specific hazard models, sub-distribution hazard models and different propensity score approaches demonstrated that different models did not change the point estimates for the outcomes (p<0.05 for MACE across all models) (**Supplementary Table 4**). Amongst patients with prior MACE, PCSK9I was associated with lower risks of MACE (HR: 0.69; 95% CI: 0.47-0.99; p=0.0469) and myocardial infarction (HR: 0.47; 95% CI: 0.27-0.80; p=0.0058). The same trend was observed upon excluding patients with less than 3-month follow-up duration, less than 3-month drug exposure, patients who died within 30 days after drug exposure, and at the top or bottom 5% of propensity score matching. Furthermore, the observed trend remained consistent amongst excluding patients with liver dysfunction. (**Supplementary Table 5**).

### Falsification analysis

Hip fracture was used as negative control outcome in the falsification analysis for the comparison between PCSK9I (**Supplementary Table 6**). The results demonstrated that amongst PCSK9I users the risk of the hip fracture after adjustment (HR: 1.42; 95% CI: 0.78-2.69, p=0.3111) was similar to the ezetimibe users.

## Discussion

In this observational cohort study, the findings demonstrated that PCSK9I usage was associated with a lower risk of MACE compared to ezetimibe usage after adjustments. However, no difference was observed for the other cardiovascular outcomes. Evolucomab demonstrated superiority compared to alirocumab in reducing the risks of MACE. After performing a falsification analysis looking at hip fracture outcomes, it did not falsify the results.

### Comparison with previous studies

The occurrence of MACE in our study for PCSK9I and ezetimibe users closely resembles the results of current literature studies, which reported an incidence of 7.95 per 1000 person-year;^25^ a study examining a Dutch cohort reported an incidence of 32.5 per 1000 person-year.^26^ The relationship between PCSK9I and the management of dyslipidaemia and atherosclerotic risks is well described.^27, 28^ Nonetheless, real-life clinical data on a direct comparison between PCSK9I and ezetimibe remains finite, and many trials studying the long-term effects of this drug are still ongoing. In the present study, PCSK9I users had reduced risks of MACE outcomes compared to ezetimibe users (HR: 0.59; 95% CI: 00.37-0.92, p=0.0203).

Current meta-analyses add evidence to suggest that PCSK9I reduced the occurrence of stroke by 25% and MI by 19% compared to their controls.^29, 30^ A meta-analysis by Imbalzaon *et al.* found that the use of evolocumab or alirocumab reduced MACE occurrence by 18% with no statistical heterogeneity amongst the eight randomised control trials that were included.^31^ This is consistent with the findings by Khan et al. that describe the protective effect of PCSK9Is and ezetimibe against stroke and non-fatal MI to be significant for high cardiovascular-risk patients but not for low or moderate-risk patients.10 To further corroborate, PCSK9I usage showed an enhanced reduction of cardiovascular events irrespective of the intensity of statin therapy for patients with acute coronary syndrome.^32^

Several randomised control trials and clinical studies supported the protective effects of PCSK9Is on the occurrence of cardiovascular events in patients with elevated risk compared to placebo.^33, 34^ The Open-Label Study of Long-Term Evaluation Against Low Density Lipoprotein Cholesterol (LDL-C) (OSLER) trial showed a lower incidence of composite adverse cardiovascular events in patients who were administered evolocumab.^35^ The ODYSSEY-

OUTCOMES (Evaluation of Cardiovascular Outcomes After an Acute Coronary Syndrome During Treatment With Alirocumab) trial underscored that means of alirocumab significantly reduced MI and all-cause mortality in patients with acute coronary syndrome compared to placebo.^36^ Contrarily, opposing evidence from a meta-analysis reveal that PCSK9I did not perform superior to ezetimibe in the prevention of MACE (Risk ratio: 0.70, 95%CI: 0.40-1.20).^11,37^ Our result demonstrated that PCSK9I was associated with lower risks of MACE amongst patients with lower levels of LDL, and the differences between PCSK9I and ezetimibe narrowed as the LDL increased. Therefore, choosing between PCSK9I or ezetimibe might require cardiovascular risk stratification.^7^ Furthermore, the lack of association between PCSK9I and ezetimibe in the existing literature could be limited by the heterogenicity amongst different types of PCSK9I.

Although both types of PCSK9I generate a net advantage against cardiovascular diseases, our results suggest that evolocumab may perform more effectively relative to alirocumab in reducing MACE risk. Furthermore, alirocumab might work better amongst patients with Q1 LDL levels, while evolocumab might work better amongst patients with Q3 or Q4 LDL levels. Previous studies reveal that the lipid-lowering effect of alirocumab is weaker compared to evolocumab.^38, 39^ Resultantly, this may have influenced the cardioprotective benefits of the drug. Alternatively, the observed discrepancy may be a result of differences in active ingredients and availability of dosage forms. However, a Spanish multicenter observational study showed similar safety profiles and no significant differences in the frequency of MACE occurrence between evolocumab and alirocumab.^40^

Our results do not necessarily mean that ezetimibe increases the risks of MACE outcomes. It is still plausible to postulate that ezetimibe may possess cardioprotective effects but may not be as potent compared to PCSK9I; however, this hypothesis requires further verification through further randomised controlled trials. In addition to its positive effect on lipid profile, multiple studies confirm the secondary cardioprotective effects and associated cost-effectiveness of ezetimibe.^41, 42^

The pathophysiological mechanisms by which PCSK9I may be related to MACE are still under investigation ^43^. PCSK9I mainly exerts its effects by inhibiting the degradation of LDL receptors and promoting uptake of cholesterol into hepatocytes, this reduces risks of atherosclerosis.^44^ In particular, evolocumab demonstrated positive effects in inducing coronary plaque regression and decreasing percent atheroma volume, thereby reducing occurrence of cardiovascular adverse events.^45^

### Clinical implications

The effects of PCSK9I and ezetimibe on reducing MACE outcomes and associated mortality have become a growing field of research interest.^46^ Current guidelines recommend PCSK9I and ezetimibe as second-line therapies for uncontrolled LDL-C, cardiovascular disease and familial hypercholesterolemia when statins are ineffective or intolerable.^47, 48^ However, the question regarding the choice between PCSK9I and ezetimibe remained open. Our result demonstrated that PCSK9I usage was associated with a lower risk of MACE and that evolocumab might be better than alirocumab, and the relationship between LDL level and the choice of drugs may provide more insights regarding the clinical decision-making process. Therefore, our results may encourage researchers to revisit clinical guidelines. Although current findings on the cardioprotective effects of PCSK9I are promising, further investigation is needed to provide more comprehensive data on the safety and efficacy.

Despite the growing body of literature on the effects of PCSK9I, there is limited knowledge regarding its real-life implications, long-term safety and cost-effectiveness.^49, 50^ The high costs of PCSK9I usage and the fact that it requires injection limits its accessibility for patients with high cardiovascular risk and may place a significant burden on clinical practices. Current studies recommend a price reduction of 60-65% in order to achieve cost-effectiveness.^51^ There have been concerns raised about high discontinuation rates and low adherence to PCSK9I. Hence, more studies need to be conducted to better understand the drug utilization patterns and optimize therapeutic efficacy.^52^

### Limitations

There are several limitations in this study that should be noted. Due to the observational nature of this study design, certain data variables such as smoking, BMI, alcohol use and family history of MACE could not be acquired and analysed. Besides, the data may also be susceptible to under-coding, missing data or other coding errors, leading to information bias. In compensation for this, comorbidities and laboratory results related to MACE were used to infer potential risk variables. Furthermore, falsification analysis was performed between PCSK9I and ezetimibe in order to reduce the risk of residual confounding. As the results did not falsify the validity of this study, this suggests that residual bias was unlikely a contributing factor to our results.

Besides, since PCSK9I remained a relatively novel drug, the sample size of the patients on PCSK9I was relatively small, such that the association between PCSK9I and MACE on myocardial infarction, stroke/ transient ischemic attack and all-cause mortality might be limited by the sample size. Furthermore, the direct comparison between evolocumab and alirocumab was insignificant due to insufficient power. However, this study already comprised all patients with PCSK9I in this locality. In the future, larger cohorts with longer follow-up duration should be built.

Notwithstanding, the retrospective design of our study indicates that all derived relationships are correlational in nature. Therefore, it is pertinent to encourage more randomised clinical trials that investigate the effect of non-statin lipid-lowering drugs on MACE as the primary outcome and identify any possible causal links.

## Conclusion

This population-based cohort study suggested that PCSK9I users were associated with lower risks of new-onset MACE amongst dyslipidaemia patients compared to ezetimibe users. Evolucomab might be better than alirocumab in terms of preventing new-onset MACE. In the future, more long-term studies are necessary to substantiate the potential effects of PCSK9I on MACE.

## Ethical approval statement

This study was approved by the Institutional Review Board of the University of Hong Kong/Hospital Authority Hong Kong West Cluster (HKU/HA HKWC IRB) (UW-20-250) and complied with the Declaration of Helsinki.

## Availability of data and materials

An anonymised version without identifiable or personal information is available from the corresponding authors upon reasonable request for research purposes.

## Guarantor Statement

All authors approved the final version of the manuscript. GT is the guarantor of this work and, as such, had full access to all the data in the study and takes responsibility for the integrity of the data and the accuracy of the data analysis.

## Author contributions

Data analysis: OHIC, LL, JZ

Data review: OHIC, LL, GT, JZ

Data acquisition: OHIC, HHHP, BKHL, AKCW

Data interpretation: CTSC, LL, QL, CC, JZ

Critical revision of manuscription: AKCW, CC, TL, GL, BMYC, GT, JZ

Supervision: BMYC, GT, JZ

Manuscript writing: OHIC, LL, CTSC, GT, JZ

Manuscript revision: OHIC, LL, QL, AKCW, TL, GL, BMYC, GT, JZ

## Acknowledgements

All the authors, and colleagues the Hospital Authority for providing de-identified clinical data are equally thanked for their contributions to this research. Special thanks to the support of the National Natural Science Foundation of China (81970270, 82170327 to TL) and the Tianjin Key Medical Discipline (Specialty) Construction Project (TJYXZDXK-029A). Structural graphical abstract and Figure 1 are created with BioRender.com.

