## Supplementary Appendix for "Comparisons of Proprotein Convertase Subtilisin/Kexin Type 9 Inhibitors (PCSK9I) versus Ezetimibe on Major Adverse Cardiovascular Events Amongst Patients with Dyslipidaemia: A Population-Based Study"

[1] Division of Clinical Pharmacology and Therapeutics, Department of Medicine, LKS Faculty of Medicine, The University of Hong Kong, Hong Kong, China
[2] Diabetes Research Unit, Cardiovascular Analytics Group, PowerHealth Limited, Hong Kong, China
[3] Department of Biostatistics & Health Informatics, Institute of Psychiatry, Psychology & Neuroscience, King's College London, London, United Kingdom
[4] Institute of Biomedical Engineering, Department of Engendering Science, University of Oxford, Oxford, United Kingdom
[5] Emergency Medicine Unit, Li Ka Shing Faculty of Medicine, The University of Hong Kong, Hong Kong, China
[6] Department of Medicine, Queen Mary Hospital, Hong Kong, China
[7] Tianjin Key Laboratory of Ionic-Molecular Function of Cardiovascular Disease, Department of Cardiology, Tianjin Institute of Cardiology, Second Hospital of Tianjin Medical University, Tianjin 300211, China
[8] Liverpool Centre for Cardiovascular Science at University of Liverpool, Liverpool John Moores University and Liverpool Heart & Chest Hospital, Liverpool, United Kingdom; and Danish Center for Health Services Research, Department of Clinical Medicine, Aalborg University, Aalborg, Denmark
[9] Kent and Medway Medical School, Canterbury, Kent, CT2 7NT, United Kingdom
[10] School of Nursing and Health Studies, Hong Kong Metropolitan University, Hong Kong, China
[11] Division of Health Science, Warwick Medical School, University of Warwick, Coventry, United Kingdom
* Contributed equally
^ Correspondence to:
Gary Tse, MD DM PhD FRCP
Kent and Medway Medical School, Canterbury, United Kingdom
Tianjin Institute of Cardiology, The Second Hospital of Tianjin Medical University, Tianjin 300211, China
School of Nursing and Health Studies, Hong Kong Metropolitan University, Hong Kong, China

Jiandong Zhou, PhD,
Division of Health Science, Warwick Medical School, University of Warwick, United Kingdom


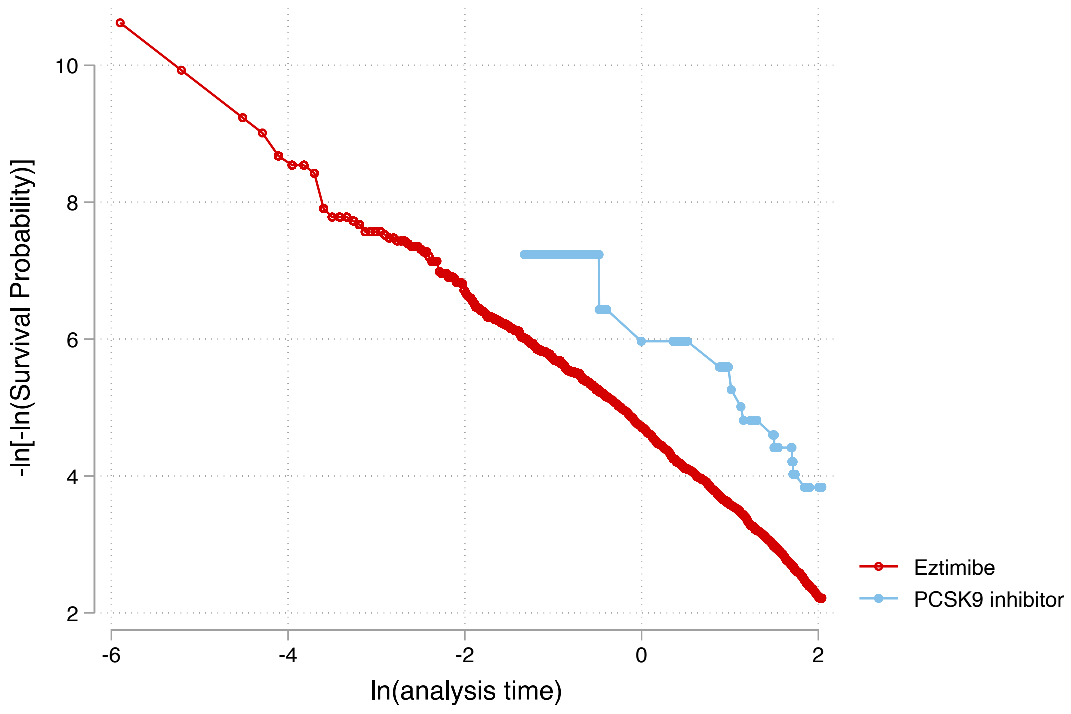

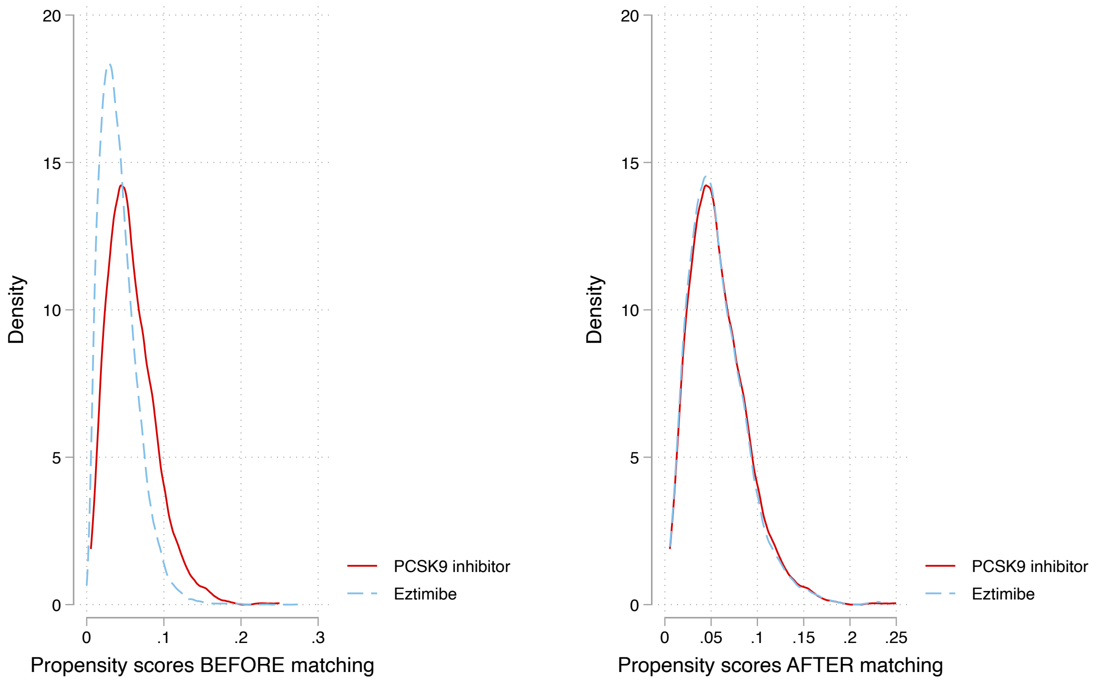

Nearest neighbor search strategy with caliper=0.1.

**Supplementary Figure 1. Propensity score matching comparisons and proportional hazard assumption testing for PCSK9 inhibitor versus ezetimibe before and after 1:3 matching with nearest neighbour search strategy with calliper of 0.1**

**Supplementary Figure 2. Cumulative incidence curves for MACE, and all-cause mortality stratified by patient gender at initial drug exposure of PCSK9 inhibitor and ezetimibe in the matched cohort**

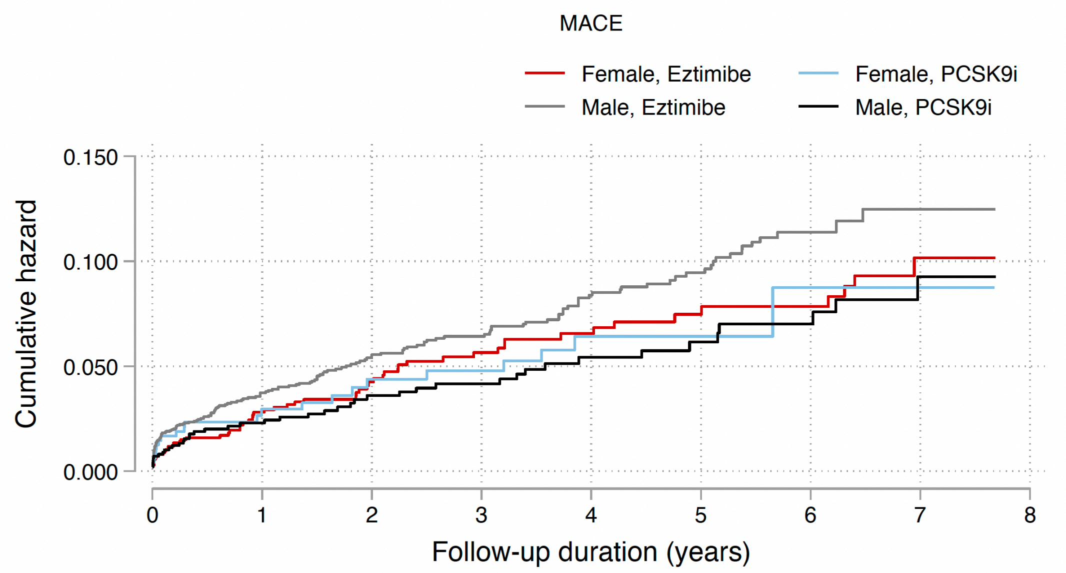

Number at risk

| | 0 | 1 | 2 | 3 | 4 | 5 | 6 | 7 | 8 |
|---|---|---|---|---|---|---|---|---|---|
| Female, Eztimibe | 1378 | 814 | 606 | 475 | 364 | 273 | 206 | 116 | 0 |
| Female, PCSK9i | 484 | 328 | 258 | 212 | 153 | 115 | 78 | 46 | 0 |
| Male, Eztimibe | 3053 | 1784 | 1332 | 1030 | 766 | 554 | 379 | 230 | 0 |
| Male, PCSK9i | 993 | 695 | 559 | 436 | 328 | 237 | 173 | 90 | 0 |

Cumulative incidence curve by drug use after propensity score matching

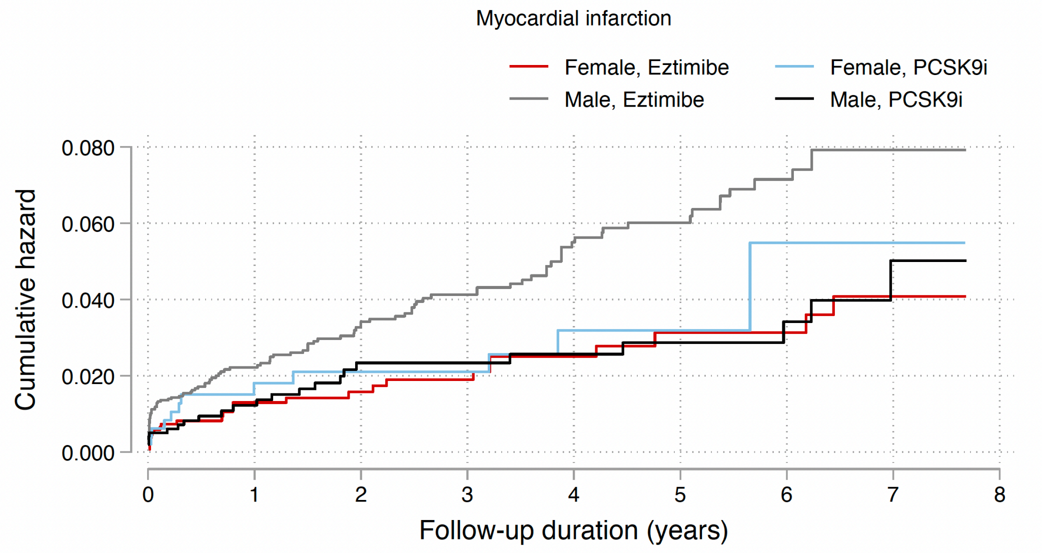

Number at risk

| | 0 | 1 | 2 | 3 | 4 | 5 | 6 | 7 | 8 |
|---|---|---|---|---|---|---|---|---|---|
| Female, Eztimibe | 1378 | 830 | 623 | 493 | 376 | 281 | 213 | 121 | 0 |
| Female, PCSK9i | 484 | 333 | 265 | 220 | 158 | 118 | 78 | 46 | 0 |
| Male, Eztimibe | 3053 | 1816 | 1362 | 1058 | 793 | 575 | 391 | 240 | 0 |
| Male, PCSK9i | 993 | 702 | 567 | 446 | 340 | 245 | 180 | 95 | 0 |

Cumulative incidence curve by drug use after propensity score matching

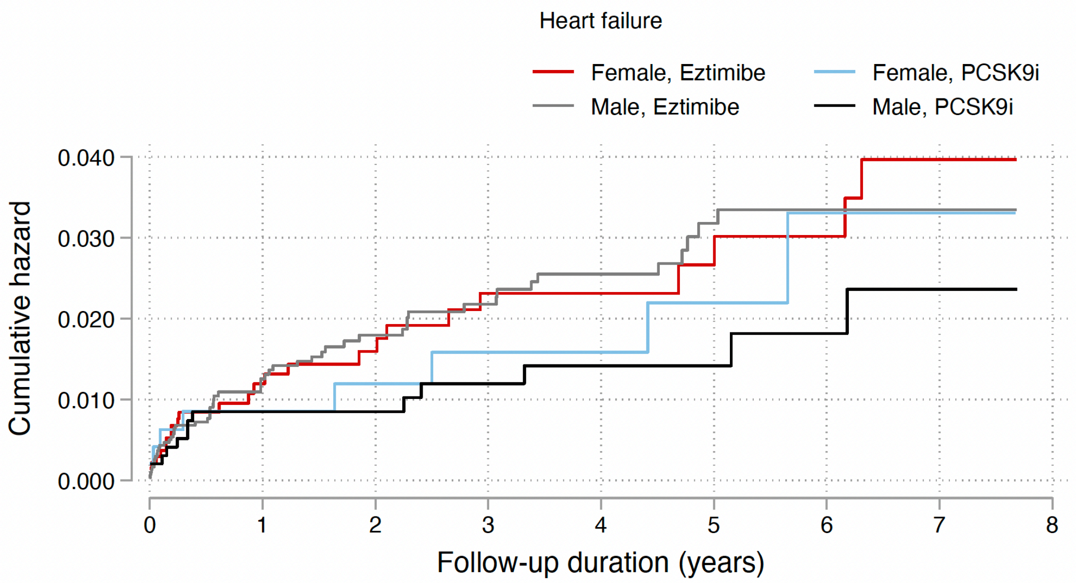

Number at risk

| | | | | | | | | | |
|---|---|---|---|---|---|---|---|---|---|
| Female, Eztimibe | 1378 | 830 | 625 | 492 | 378 | 284 | 211 | 117 | 0 |
| Female, PCSK9i | 484 | 337 | 268 | 221 | 164 | 122 | 82 | 50 | 0 |
| Male, Eztimibe | 3053 | 1841 | 1382 | 1081 | 818 | 598 | 416 | 256 | 0 |
| Male, PCSK9i | 993 | 708 | 578 | 453 | 346 | 250 | 183 | 95 | 0 |

Cumulative incidence curve by drug use after propensity score matching

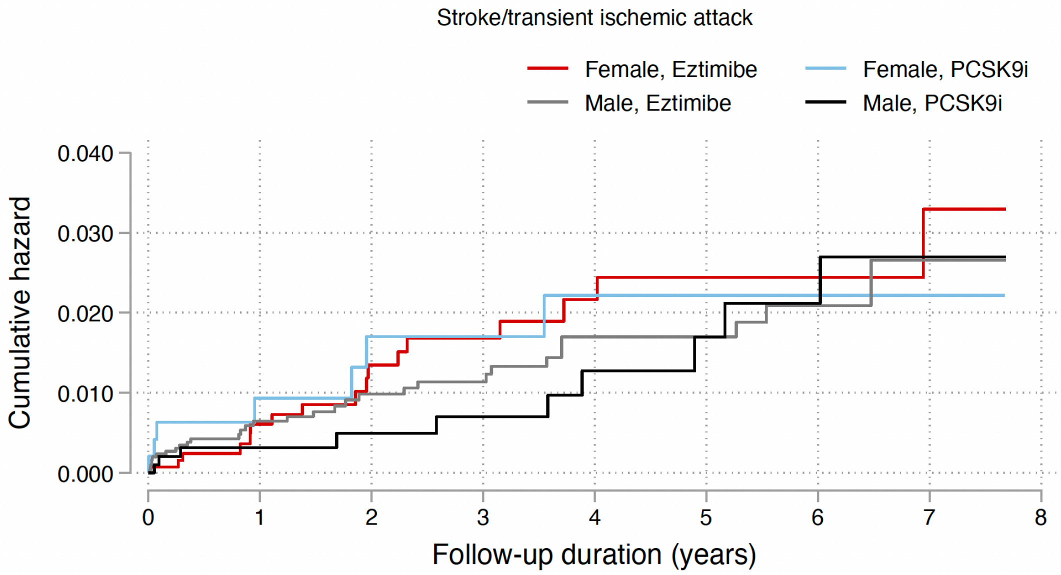

Number at risk

| | | | | | | | | | |
|---|---|---|---|---|---|---|---|---|---|
| Female, Eztimibe | 1378 | 814 | 606 | 475 | 364 | 273 | 206 | 116 | 0 |
| Female, PCSK9i | 484 | 328 | 258 | 212 | 153 | 115 | 78 | 46 | 0 |
| Male, Eztimibe | 3053 | 1785 | 1334 | 1031 | 766 | 554 | 379 | 230 | 0 |
| Male, PCSK9i | 993 | 695 | 559 | 436 | 328 | 237 | 173 | 90 | 0 |

Cumulative incidence curve by drug use after propensity score matching

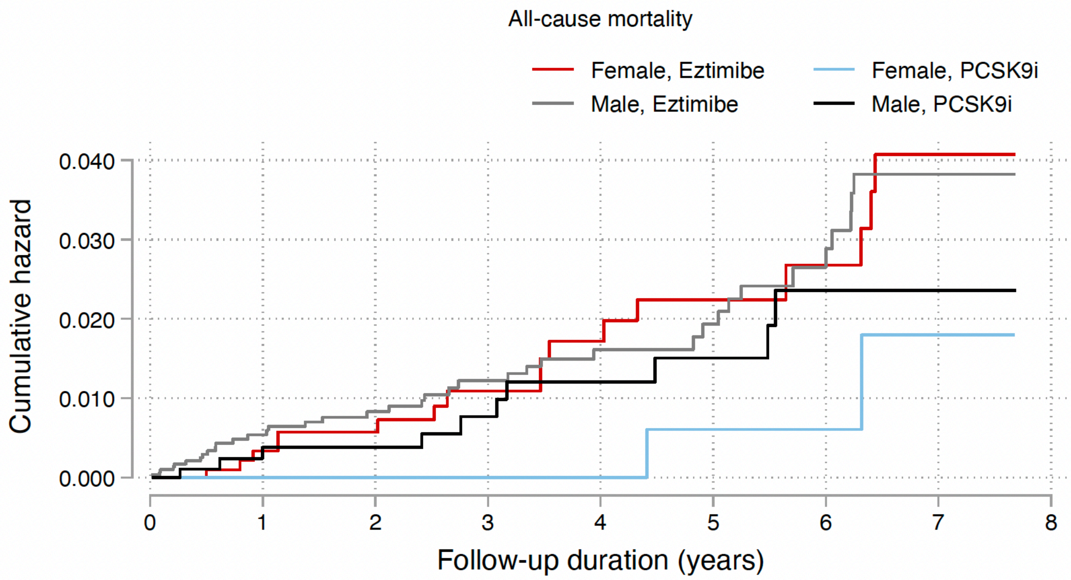

Number at risk

| | | | | | | | | | |
|---|---|---|---|---|---|---|---|---|---|
| Female, Eztimibe | 1378 | 842 | 635 | 503 | 384 | 290 | 216 | 122 | 0 |
| Female, PCSK9i | 484 | 341 | 273 | 225 | 165 | 123 | 84 | 51 | 0 |
| Male, Eztimibe | 3053 | 1868 | 1411 | 1109 | 846 | 622 | 428 | 263 | 0 |
| Male, PCSK9i | 993 | 713 | 583 | 458 | 351 | 253 | 185 | 98 | 0 |

Cumulative incidence curve by drug use after propensity score matching

**Supplementary figure 3. Marginal effects of time weighted mean LDL, number of prior lipid-lowering drugs, and dyslipidaemia duration with 95% CIs on adverse outcomes by drug use in the matched cohort.**

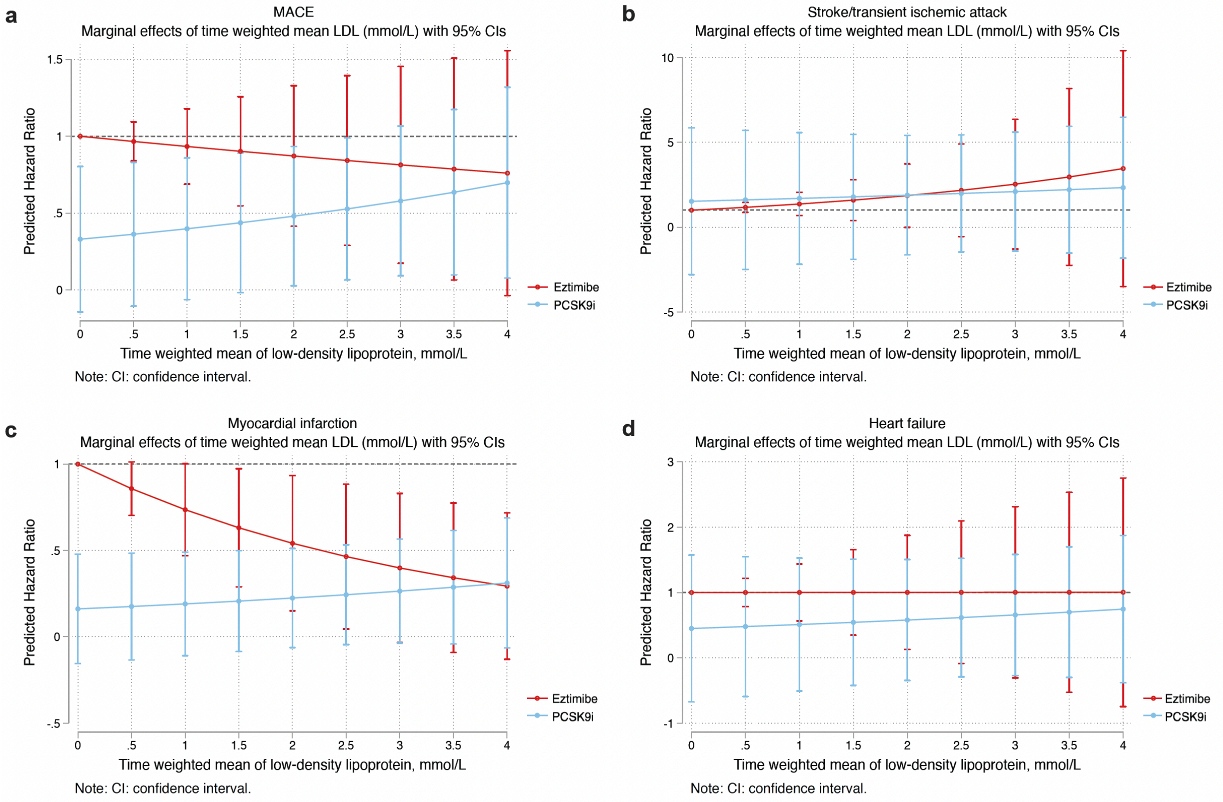

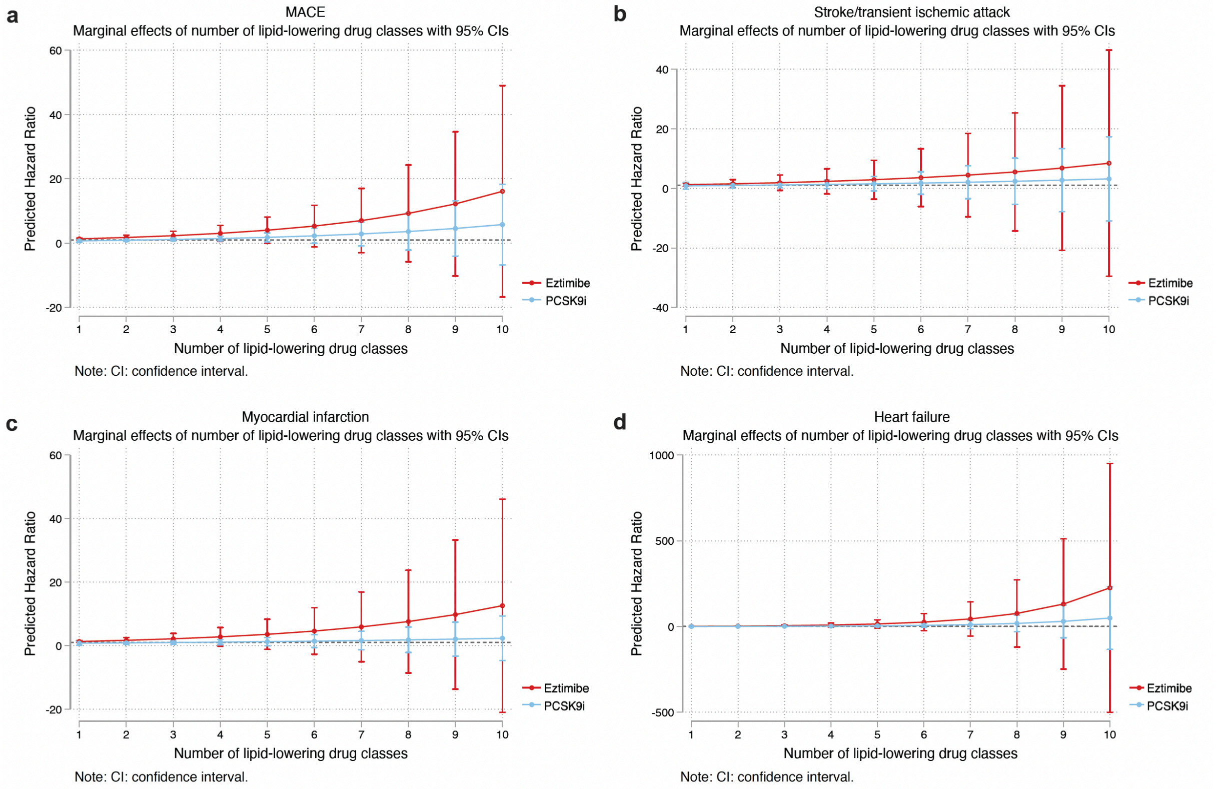

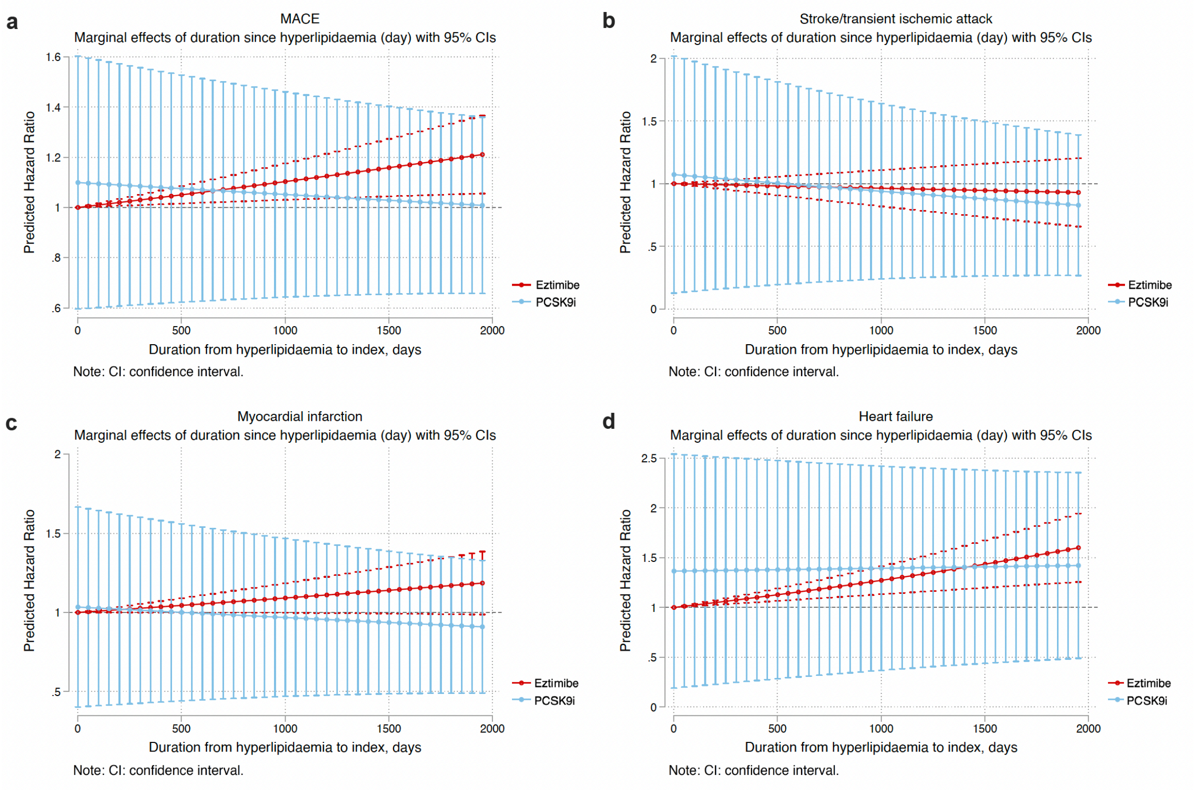
a
MACE
Marginal effects of duration since hyperlipidaemia (day) with 95% CIs
Predicted Hazard Ratio
Duration from hyperlipidaemia to index, days
Eztimibe
PCSK9i
Note: CI: confidence interval.
b
Stroke/transient ischemic attack
Marginal effects of duration since hyperlipidaemia (day) with 95% CIs
Predicted Hazard Ratio
Duration from hyperlipidaemia to index, days
Eztimibe
PCSK9i
Note: CI: confidence interval.
c
Myocardial infarction
Marginal effects of duration since hyperlipidaemia (day) with 95% CIs
Predicted Hazard Ratio
Duration from hyperlipidaemia to index, days
Eztimibe
PCSK9i
Note: CI: confidence interval.
d
Heart failure
Marginal effects of duration since hyperlipidaemia (day) with 95% CIs
Predicted Hazard Ratio
Duration from hyperlipidaemia to index, days
Eztimibe
PCSK9i
Note: CI: confidence interval.

**Supplementary Table 1. The International Classification of Diseases, Clinical Modification (ICD-9) and (ICD-10) codes for definitions of past comorbidities and outcomes.**

| **Adverse outcome of interest** |
|---|
| MACE:<br>**1.** cardiovascular mortality<br>**2.** myocardial infarction<br>**3.** heart failure<br>**4.** stroke/ transient ischaemic attack |
| **Cardiovascular mortality:** ICD-10: I00-I78 |
| **Acute myocardial infarction:** 410 410.01 410.02 410.1 410.11 410.12 410.2 410.21 410.22 410.3 410.31 410.32 410.4 410.41 410.42 410.5 410.51 410.52 410.6 410.61 410.62 410.7 410.71 410.72 410.8 410.81 410.82 410.9 410.91 410.92 |
| **Heart failure:** 428 428 428.1 428.2 428.2 428.21 428.22 428.23 428.3 428.3 428.31 428.32 428.33 428.4 428.4 428.41 428.42 428.43 428.9 398.91 402.01 402.11 402.91 404.01 404.03 404.11 404.13 404.91 404.93 |
| **Stroke/transient ischemic attack:** 435 435.1 435.2 435.3 435.8 435.9 433.81 433.91 434 436 437 437.1 433.31 433.01 434.01 434.1 434.11 434.9 434.91 437.2 437.3 437.4 437.5 437.6 437.7 437.8 437.9 430 431 432 432.1 432.9 |
| **Past comorbidities** |
| **Cancer**: 140-239 |
| **Hypertension**: 401 401.1 401.9 402 402.01 402.1 402.11 402.9 402.91 403 403.01 403.1 403.11 403.9 403.91 404 404.01 404.02 404.03 404.1 404.11 404.12 404.13 404.9 404.91 404.92 404.93 405 405.01 405.09 405.1 405.11 405.19 405.9 405.91 405.99 437.2 + history of uses of anti-hypertensives |
| **Hyperlipidaemia**: 272.0 272.1 272.2 272.3 272.4 + history of uses of lipid-lowering drugs + lipid laboratory results |
| **Liver disease:** 275.1 275.0 572.0 572.4 572.1 572.3 572.8 573.0 573.4 573.8 573.9 |
| **Gastrointestinal bleeding:** 578.9 |
| **Atrial fibrillation:** 427.31 429.4 |
| **Ischemic heart disease:** 410.01 410.02 410.1 410.11 410.12 410.2 410.21 410.22 410.3 410.31 410.32 410.4 410.41 410.42 410.5 410.51 410.52 410.6 410.61 410.62 410.7 410.71 410.72 410.8 410.81 410.82 410.9 410.91 410.92 411 411.1 411.8 411.81 411.89 413 413.1 413.9 414 414.01 414.02 414.03 414.04 414.05 414.06 414.07 414.1 414.11 414.12 414.19 414.2 414.3 414.4 414.8 414.9 410 412 |
| **Renal diseases:** 582 582 582.1 582.2 582.4 582.8 582.81 582.89 582.9 583 583 583.1 583.2 583.4 583.6 583.7 585 585.1 585.2 585.3 585.4 585.5 585.6 585.9 586 588 588 588.1 588.8 588.81 588.89 588.9 |
| **Diabetic retinopathy:** 250.5 361 362.01 362.02 362.1 362.53 362.81 362.82 362.83 369 379.23 |
| **Diabetic nephropathy:** 250.40 250.41 250.42 250.43 |
| **Diabetic neuropathy**: 250.6, 337.0, 337.1, 354.0 – 355.9, 356.9, 357.2, 358.1, 536.3, 564.5, 596.54, 713.5, 951.0, 951.1, 951.3 |
| **Chronic obstructive pulmonary disease** 491.0 491.1 491.2 491.8 491.9 492.0 492.8 496 |
| **Venous thromboembolism:** 415.1 451, 453.1 453.2 453.3 453.8 453.9 |
| **Hip fracture:** 733.14 733.15 733.81 733.82 733.96 808 808.1 820 820.01 820.02 820.03 820.09 820.10 820.11 820.12 820.13 820.19 820.20 820.21 820.22 820.30 820.31 820.32 820.8 820.9 |

**Supplementary table 2. Multivariate Cox regression models with adjustments to predict adverse outcomes in the matched cohort.**

* for p≤ 0.05, ** for p ≤ 0.01, *** for p ≤ 0.001; HR: hazard ratio; CI: confidence interval;

**Model 1** adjusted for significant demographics.

**Model 2** adjusted for significant demographics, and past comorbidities.

**Model 3** adjusted for significant demographics, past comorbidities, duration from earliest diabetes mellitus date to initial drug exposure date, and number of prior hospitalizations.

**Model 4** adjusted for significant demographics, past comorbidities, duration of diabetes mellitus, and number of prior hospitalizations, number of anti-diabetic drugs, and non-PCSK9 inhibitor/ezetimibe medications.

**Model 5** adjusted for significant demographics, past comorbidities, duration of diabetes mellitus, and number of prior hospitalizations, number of anti-diabetic drugs, non-PCSK9 inhibitor/ezetimibe medications.

**Model 6** adjusted for significant demographics, past comorbidities, duration of diabetes mellitus, and number of prior hospitalizations, number of anti-diabetic drugs, non-PCSK9 inhibitor/ezetimibe medications, abbreviated MDRD, HbA1c, fasting glucose.

**Model 7** adjusted for significant demographics, past comorbidities, duration of diabetes mellitus, and number of prior hospitalizations, number of anti-diabetic drugs, non-PCSK9 inhibitor/ezetimibe medications, abbreviated MDRD, and time-weighted means of lipid and glucose tests.

| **Model** | **All-cause mortality HR [95% CI];P value** | **MACE HR [95% CI];P value** | **Myocardial infarction HR [95% CI];P value** | **Heart failure HR [95% CI];P value** | **Stroke/transient ischemic attack HR [95% CI];P value** |
|---|---|---|---|---|---|
| Model 1 | 0.32[0.15-0.73];0.0017** | 0.61[0.42-0.89];0.0002*** | 0.55[0.40-0.89];0.0011** | 0.50[0.31-0.82];0.0018** | 0.87[0.50-1.52];0.6360 |
| Model 2 | 0.34[0.16-0.69];0.0029** | 0.65[0.49-0.86];0.0027** | 0.62[0.43-0.91];0.0135* | 0.51[0.30-0.86];0.0112* | 0.72[0.40-1.28];0.2591 |
| Model 3 | 0.47[0.24-0.94];0.0321* | 0.67[0.51-0.89];0.0056** | 0.63[0.43-0.92];0.0155* | 0.54[0.32-0.90];0.0180* | 0.79[0.45-1.39];0.4100 |
| Model 4 | 0.60[0.30-1.19];0.1419 | 0.67[0.50-0.89];0.0054** | 0.63[0.43-0.93];0.0185* | 0.68[0.41-1.14];0.1431 | 0.78[0.43-1.41];0.4083 |
| Model 5 | 0.51[0.22-1.18];0.1142 | 0.59[0.38-0.90];0.0140* | 0.60[0.33-1.08];0.0896 | 0.85[0.447-1.602];0.6077 | 0.48[0.17-1.31];0.1515 |
| Model 6 | 0.39[0.15-1.02];0.0550 | 0.54[0.35-0.84];0.0065** | 0.55[0.29-1.04];0.0671 | 0.85[0.438-1.610];0.5989 | 0.49[0.17-1.38];0.1774 |
| Model 7 | 0.45[0.17-1.20];0.1101 | 0.59[0.37-0.92];0.0203* | 0.55[0.28-1.08];0.0828 | 0.89[0.455-1.756];0.7459 | 0.50[0.18-1.41];0.1917 |

**Supplementary Table 3. Number of adverse outcomes and time of the PCSK9 inhibitor subtypes to adverse outcomes in the matched cohort.**

| Characteristics | Only alirocumab (N=555) Mean(SD);N or Count(%) | Only evolocumab (N=110) Mean(SD);N or Count(%) | Eztimibe (N=4431) Mean(SD);N or Count(%) |
|---|---|---|---|
| All-cause mortality | 9(1.62%) | 0(0.00%) | 55(1.24%) |
| Time to mortality, years | 3.1(2.5);n=555 | 3.2(2.4);n=110 | 2.6(2.5);n=4431 |
| MACE | 49(8.82%) | 5(4.54%) | 235(5.30%) |
| Time to MACE, years | 2.9(2.5);n=555 | 3.0(2.4);n=110 | 2.5(2.4);n=4431 |
| Myocardial infarction | 27(4.86%) | 3(2.72%) | 139(3.13%) |
| Time to MI, years | 3.0(2.5);n=555 | 3.1(2.4);n=110 | 2.6(2.4);n=4431 |
| Heart failure after drug use | 13(2.34%) | 3(2.72%) | 80(1.80%) |
| Time to HF, years | 3.1(2.5);n=555 | 3.1(2.5);n=110 | 2.6(2.4);n=4431 |
| Stroke/transient ischemic attack | 13(2.34%) | 0(0.00%) | 49(1.10%) |
| Time to Stroke/TIA, years | 2.9(2.5);n=555 | 3.0(2.4);n=110 | 2.5(2.4);n=4431 |

**Supplementary Table 4. Sensitivity analyses for exposure effects of PCSK9 inhibitor v.s. ezetimibe on diverse outcomes using different models.**

* for p≤ 0.05, ** for p ≤ 0.01, *** for p ≤ 0.001; HR: hazard ratio; CI: confidence interval; PS: propensity score; IPTW: inverse probability of treatment weighting, SIPTW: stable inverse probability of treatment weighting.

| **Model** | **All-cause mortality HR [95% CI];P value** | **MACE HR [95% CI];P value** | **Myocardial infarction HR [95% CI];P value** | **Heart failure HR [95% CI];P value** | **Stroke/transient ischemic attack HR [95% CI];P value** |
|---|---|---|---|---|---|
| Cause-specific hazard models | 0.48[0.22-1.05];0.1004 | 0.53[0.34-0.85];0.0014** | 0.56[0.29-1.11];0.0913 | 0.79[0.32-1.56];0.5611 | 0.44[0.34-1.34];0.2322 |
| Sub-distribution hazard models | 0.51[0.13-1.01];0.1204 | 0.57[0.29-0.92];0.0004*** | 0.52[0.34-1.09];0.1109 | 0.82[0.44-1.41];0.4511 | 0.55[0.32-1.32];0.4311 |
| PS stratification | 0.46[0.12-1.13];0.1112 | 0.65[0.36-0.91];0.0015** | 0.65[0.25-1.24];0.2131 | 0.83[0.36-1.44];0.3511 | 0.66[0.45-1.52];0.4212 |
| PS with IPTW | 0.49[0.25-1.32];0.1604 | 0.56[0.25-0.88];0.0101** | 0.56[0.18-1.22];0.1311 | 0.86[0.48-1.89];0.4551 | 0.69[0.32-1.72];0.2504 |
| PS with SIPTW | 0.53[0.34-1.29];0.1523 | 0.66[0.34-0.85];0.0107* | 0.59[0.22-1.24];0.1421 | 0.91[0.69-1.81];0.7822 | 0.59[0.25-1.56];0.5521 |

**Supplementary Table 5. Sensitivity analysis of PCSK9 inhibitor v.s. ezetimibe on diverse outcomes.**

Adjusted for significant demographics, past comorbidities, duration of diabetes mellitus, and number of prior hospitalizations, number of anti-diabetic drugs, non-PCSK9 inhibitor/ezetimibe medications, abbreviated MDRD, and time-weighted means of lipid and glucose tests.

| **Model** | **All-cause mortality HR [95% CI];P value** | **MACE HR [95% CI];P value** | **Myocardial infarction HR [95% CI];P value** | **Heart failure H R [95% CI];P value** | **Stroke/transient ischemic attack HR [95% CI];P value** |
|---|---|---|---|---|---|
| Only patients with prior MACE | 0.59[0.26-1.35];0.2122 | 0.69[0.47-0.99];0.0469* | 0.47[0.27-0.80];0.0058** | 0.72[0.40-1.30];0.2781 | 0.79[0.36-1.77];0.5698 |
| Exclude patients with prior MACE | 0.43[0.17-1.14];0.0892 | 0.80[0.53-1.20];0.2834 | 1.04[0.62-1.76];0.8816 | 0.61[0.26-1.40];0.2404 | 0.88[0.41-1.91];0.7552 |
| Exclude patients with less than 3-month follow-up duration | 0.55[0.29-1.04];0.0663 | 0.74[0.56-0.97];0.0283* | 0.68[0.47-0.98];0.0404* | 0.68[0.42-1.11];0.1237 | 0.84[0.48-1.46];0.5249 |
| Exclude patients with less than 3-month drug exposure | 0.51[0.26-0.97];0.0412* | 0.73[0.56-0.97];0.0270* | 0.68[0.47-0.98];0.0391* | 0.68[0.42-1.11];0.1210 | 0.83[0.48-1.45];0.5209 |
| Exclude patients who died within 30 | 0.52[0.28-0.97];0.0390* | 0.74[0.56-0.97];0.0308* | 0.68[0.47-0.99];0.0416* | 0.69[0.42-1.12];0.1322 | 0.84[0.48-1.46];0.5393 |

| | | | | | |
|---|---|---|---|---|---|
| days after drug exposure | | | | | |
| Exclude patients with liver dysfunction defined as AST or ALT >3× ULN | 0.41[0.20-0.83];0.0137* | 0.70[0.53-0.93];0.0153* | 0.65[0.44-0.95];0.0261* | 0.61[0.36-1.04];0.0701 | 0.79[0.44-1.42];0.4286 |
| Exclude patients at the top or bottom 5% of propensity score matching | 0.53[0.28-1.00];0.0508 | 0.74[0.56-0.97];0.0315* | 0.68[0.47-0.99];0.0438* | 0.69[0.42-1.12];0.1295 | 0.84[0.48-1.46];0.5341 |

**Supplementary Table 6. Falsification analysis: Exposure effects of PCSK9 inhibitor versus ezetimibe on new onset hip fracture in the matched cohort after 1:3 propensity score matching.**

* for p≤ 0.05, ** for p ≤ 0.01, *** for p ≤ 0.001; HR: hazard ratio; CI: confidence interval

Adjusted for significant demographics, past comorbidities, duration of diabetes mellitus, and number of prior hospitalizations, number of anti-diabetic drugs, non-PCSK9 inhibitor/ezetimibe medications, abbreviated MDRD, and time-weighted means of lipid and glucose tests.

| | **Number of hip fracture events Count(%)** | **Time to hip fracture, days (IQR)** | **Hip fracture after drug use HR[95% CI];P value** |
|---|---|---|---|
| Ezetimibe (N=4431) | 28(0.68%) | 582(180-1645) | 1 [Reference] |
| PCSK9 inhibitor (N=1477) | 17(1.15%) | 942(230-1985) | 1.42[0.78-2.69];0.3111 |
| Alirocumab (N=1367) | 13(0.95%) | 943(223-1986) | 1.45[0.89-2.98];0.3019 |
| Evolocumab (N=922) | 5(0.54%) | 935(232-1985) | 0.96[0.55-2.15];0.8155 |
